# Neuromechanical Predictors of Clinical Scores of Balance and Functional Mobility in Chronic Stroke Survivors – A Machine Learning Approach

**DOI:** 10.1101/2025.10.19.25338257

**Authors:** Komal K. Kukkar, Sheng Li, Irving Weinberg, Pranav J. Parikh

## Abstract

Clinical tests such as the Berg Balance Scale (BBS) and Timed Up and Go (TUG) are used to assess balance and functional mobility following stroke. These tests are subjective due to dependence on the assessor’s judgment and the patient’s effort, potentially affecting their sensitivity to early or subtle balance recovery. This study aimed to identify objective markers of BBS and TUG among corticomuscular coherence (CMC) and force platform center of pressure (COP) measures using machine learning in 18 chronic stroke patients and 15 age-matched healthy adults. Participants performed a continuous balance task on a sway-referenced force platform with simultaneous recording of EEG, EMG, and COP data. We used a two-stage machine learning approach: first, a binary classifier, such as XGBoost and Elastic Net models, reduced dimensionality while preserving interpretability, and selected features that differentiated between stroke and healthy controls; second, regression models used selected features to identify predictors of BBS and TUG. Tibialis anterior delta-band CMC and medio-lateral root mean square COP predicted BBS and TUG. These features capture the dynamic stability mechanisms shared across the two clinical tests of balance and functional mobility. Soleus theta-band CMC asymmetry index predicted BBS, whereas TUG was predicted by biceps femoris beta-band CMC asymmetry index and rectus femoris beta-band CMC. These muscle-specific measures highlighted the use of ankle and hip strategies for the control of balance during BBS and TUG tests, respectively. Our study provides objective neurophysiological and biomechanical markers that may be sensitive to subtle changes in balance following stroke.

## INTRODUCTION

Up to 73% of stroke survivors fall within 6-12 months post-discharge, causing a decline in activities of daily living (ADLs), independence, and quality of life [1–4]. Poor balance and functional mobility put a stroke patient at a greater risk for falls [5–7]. Assessment of balance and functional mobility provides knowledge of an individual’s ability to maintain stability, predict fall risk, track balance recovery after stroke, and tailor interventions to improve function [8]. Balance and functional mobility are commonly assessed using clinical tests such as the Berg Balance Scale (BBS) and the Timed Up and Go (TUG) test [9,10]. Although widely used, BBS and TUG tests are known to have limitations. BBS and TUG are prone to subjectivity because the ratings can be influenced by the assessor’s judgment and the patient’s effort [11,12]. Additionally, BBS can exhibit a ceiling effect in stroke patients [11]. These limitations reduce their sensitivity to early or subtle recovery. There is a need for objective, non-invasive markers that can accurately predict clinical outcomes, provide greater sensitivity, and minimize reliance on subjective ratings.

Several studies have investigated neurophysiological measures such as electroencephalography (EEG), electromyography (EMG), corticomuscular coherence (CMC), and force platform-based center of pressure (COP) metrics as potential predictors of balance and functional mobility impairments in stroke survivors. These measures are suggested to have a greater sensitivity to balance changes with intervention than traditional clinical assessments [13,14]. Mainly, cortico-muscular coherence is studied to quantify the degree of oscillatory coupling (i.e., in the frequency domain) between the EEG and EMG signals [13]. Reduced beta-band CMC has been linked to impaired corticomuscular communication and longer TUG times during functional mobility in stroke participants [15,16], while lower-limb beta-CMC correlates with walking stability [17,18]. Reduced delta-band CMC, when the somatosensory contribution to the dynamic balance is compromised using sway-referencing, during standing, has been linked to poor BBS and TUG outcomes [13,19,20]. However, most neurophysiological and EMG-based studies emphasize unilateral assessments, typically focusing on the affected limb in stroke patients, overlooking bilateral interactions that are critical for understanding compensatory strategies and recovery. Evidence suggests that muscle activity from both limbs is associated with BBS and TUG outcomes [21,22]. Furthermore, task-specific and technology-assisted interventions—such as electrical stimulation, constraint-induced movement therapy, and structured exercise programs— designed to involve both limbs than just the affected side have demonstrated significant improvements in balance and functional mobility [23,24]. These findings highlight the importance of considering inter-limb interactions when assessing balance recovery in stroke patients.

Force platforms have also been used to obtain quantitative measures of balance control. COP sway obtained from an instrumented support surface differentiates high-versus low-fall-risk individuals with greater sway displacement and velocity in fall-prone groups, and classifies stroke participants in comparison to age-matched healthy controls. Increased COP sway predicts lower BBS scores and prolonged TUG times, suggesting its clinical utility in assessing balance and functional mobility limitations [25–31]. Postural sway metrics obtained during quiet standing show reductions as recovery advances [32], correlating with improved BBS and faster TUG times [33]. However, not all COP based measures correlate with BBS and TUG [34].

To the best of our knowledge, none of the previous studies investigated which measures among several CMC-based and force platform-based measures, obtained during laboratory balance tasks, best predict the clinical score of balance and functional mobility. This requires a direct comparison of potential markers to elucidate the most important parameter that predicts BBS and TUG.

Therefore, the primary aim of this study was to integrate multimodal features, encompassing force platform biomechanical measures and neurophysiological corticomuscular coherence measures using machine learning, to predict performance on the Berg Balance Scale (BBS) and Timed Up and Go (TUG) in individuals with stroke and their age-range matched healthy adults. We adopted a two-stage machine learning approach. In the first stage, binary classification models (stroke versus healthy) were applied to perform feature selection. This step identified variables that best discriminate between groups. Classification-based feature selection has been widely recommended in machine learning to reduce dimensionality while preserving interpretability [35,36]. In the second stage, the selected features were entered into regression models to predict BBS and TUG scores, ensuring that the subsequent analyses are based on clinically meaningful and statistically robust variables. The findings of this study are meant to identify clinically meaningful markers that can track recovery trajectories, improve risk stratification, and support personalized rehabilitation planning.

## METHODS

### Participants

Eighteen stroke participants (60.9 ± 9.17 years, mean ± SD; range: 37-70) and fifteen healthy adult participants (57.2 ± 7.98 years, mean ± SD; range: 35-70) provided informed written consent to participate in this research study. Demographics are presented in **Table 1**. The inclusion criteria for participants with stroke included middle cerebral artery (MCA) stroke for the first time, at least 12 months post stroke, ability to stand for 5 minutes independently without assistance, between 18-90 years of age, have MoCA score ≥ 26 [37,38], and absence of other neurological/ musculoskeletal impairments. Healthy adults (controls) were recruited if they had no history/symptoms of neurological/neuromuscular disorders affecting lower limbs with a MoCA score of 26 or greater. Healthy adults were right-foot dominant based on self-reporting of the leg with which they preferred to take a step. This study was approved by the Institutional Review Board at the University of Houston.

**Table 1.**
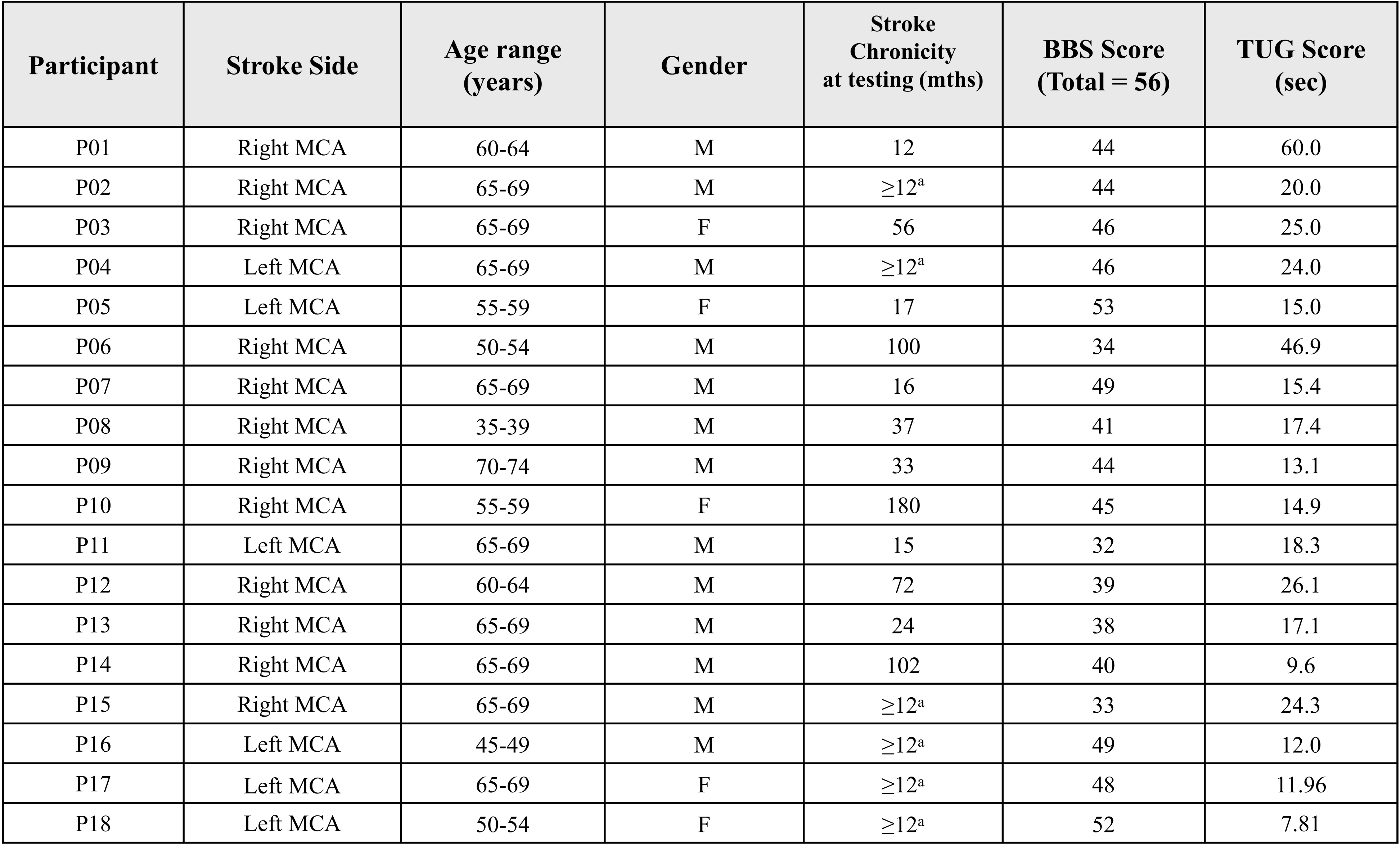
Clinical information of stroke participants. ^a^ exact data is not available; F, female; M, male; MCA, middle cerebral artery; BBS: Berg Balance Scale; TUG: Timed Up and Go.

| Participant | Stroke Side | Age range (years) | Gender | Stroke Chronicity at testing (mths) | BBS Score (Total = 56) | TUG Score (sec) |
| --- | --- | --- | --- | --- | --- | --- |
| P01 | Right MCA | 60-64 | M | 12 | 44 | 60.0 |
| P02 | Right MCA | 65-69 | M | ≥12<sup>a</sup> | 44 | 20.0 |
| P03 | Right MCA | 65-69 | F | 56 | 46 | 25.0 |
| P04 | Left MCA | 65-69 | M | ≥12<sup>a</sup> | 46 | 24.0 |
| P05 | Left MCA | 55-59 | F | 17 | 53 | 15.0 |
| P06 | Right MCA | 50-54 | M | 100 | 34 | 46.9 |
| P07 | Right MCA | 65-69 | M | 16 | 49 | 15.4 |
| P08 | Right MCA | 35-39 | M | 37 | 41 | 17.4 |
| P09 | Right MCA | 70-74 | M | 33 | 44 | 13.1 |
| P10 | Right MCA | 55-59 | F | 180 | 45 | 14.9 |
| P11 | Left MCA | 65-69 | M | 15 | 32 | 18.3 |
| P12 | Right MCA | 60-64 | M | 72 | 39 | 26.1 |
| P13 | Right MCA | 65-69 | M | 24 | 38 | 17.1 |
| P14 | Right MCA | 65-69 | M | 102 | 40 | 9.6 |
| P15 | Right MCA | 65-69 | M | ≥12<sup>a</sup> | 33 | 24.3 |
| P16 | Left MCA | 45-49 | M | ≥12<sup>a</sup> | 49 | 12.0 |
| P17 | Left MCA | 65-69 | F | ≥12<sup>a</sup> | 48 | 11.96 |
| P18 | Left MCA | 50-54 | F | ≥12<sup>a</sup> | 52 | 7.81 |

### Instrumentation

#### Computerized dynamic posturography (CDP)

A commercially available CDP Force platform (Neurocom Balance Master, Natus Medical Incorporated, Pleasanton, CA) was used to assess dynamic balance stability. It is extensively used both in clinical [39] and research settings [40] for monitoring sensory and motor performance aspects of the balance control system. The platform is equipped with a motorized dual force plate system (45.72 cm x 45.72 cm), in which ground reaction forces (GRF) from under the subject’s feet are collected by normal and shear force transducers embedded within the force plate (support surface). The system can be used to adjust the orientation of the force plate with respect to the gravitational vertical by rotating it in the sagittal plane about an axis through the subject’s ankle joint in some proportion (a pre-selected gain between −2 to +2) to the postural sway of the subject. A negative sway gain means the movement of the plate will be in the opposite direction of the subject’s COP, and a positive sway gain means the movement of the plate will be in the same direction as the subject’s COP. The transducer data were collected at 100 Hz and processed by pre-installed software on a Windows-based desktop connected to the Neurocom Balance Master (Research module, Neurocom software version 8.0, Natus Medical Incorporated, Pleasanton, CA). The Neurocom system also generated an analog timing signal, which was used to synchronize the electroencephalography (EEG) and the electromyography (EMG) system with GRF data.

### Clinical Assessment of balance and functional mobility

Berg Balance Scale (BBS): This scale is extensively used in clinical and research settings to assess static and dynamic balance. It consists of a series of 14 predetermined tasks, and each task is scored, ranging from 0-4; “0” indicates the lowest functional level, and “4” indicates the highest functional level. The score can take a value between 0 and 56, with higher scores suggestive of better balance [41].

Timed Up and Go (TUG) Test: The TUG test measures dynamic balance and functional mobility. The subjects were instructed to rise from an armchair, walk 3 meters from the marker, turn around, walk back to the chair, and sit down. The time to complete this test was measured in seconds. More time taken is associated with poor dynamic balance/functional mobility [42]. Three trials were performed, and a mean score (seconds) was used in subsequent analysis.

### Laboratory-based assessment of reactive balance control

To study cortical involvement in dynamic balance control, we customized a continuous balance task to design a dynamic balance paradigm. The balance control system must continuously process information from multisensory systems to assess balance stability and orientation of the body and initiate appropriate behavioral responses in case of disturbance to prevent a fall. During the continuous balance task, participants stood on the balance master with eyes closed while EEG, EMG, and GRF data were collected (**Fig. 1a**). The balance context was changed by varying the responsiveness of the support surface in proportion to the estimated center of mass sway – fixed or sway-referenced. The sway-referencing of the support surface reduces the contribution of lower limb somatosensory receptors to the control of dynamic balance [43] (**Fig. 1b**) and thereby manipulates the reliance on somatosensory inputs for the control of ongoing stance and modifies the feedback relationship within the balance control loop. The effects of such manipulation on dynamic stability are related to the action of the participant. We altered the relationships between postural sway and somatosensory inputs by varying the gain of the support surface in the sagittal plane in different proportions to the estimated instantaneous COM sway angle. We used a range of gains (−1.0, −0.4, 0, 0.4, 0.6, 1.0, 2.0) in the balance task of three different levels of balance difficulty: *low*, *medium,* and *high* (**Fig. 1c**). The lowest gain value of 0 (a quiet stance, fixed support surface with no sway referencing) may not be challenging to subjects and therefore classified as *low* difficulty. Higher gain values of 0.4, 0.6, and 1.0 were classified as *medium* difficulty, as subjects were expected to exhibit greater balance instability. However, the gain of 2.0, or negative gains of −0.4 and −1.0, were classified as that of *high* difficulty due to expectations that participants would have a further increase in balance instability, as done in our previous work [44–46].

**Fig 1.**
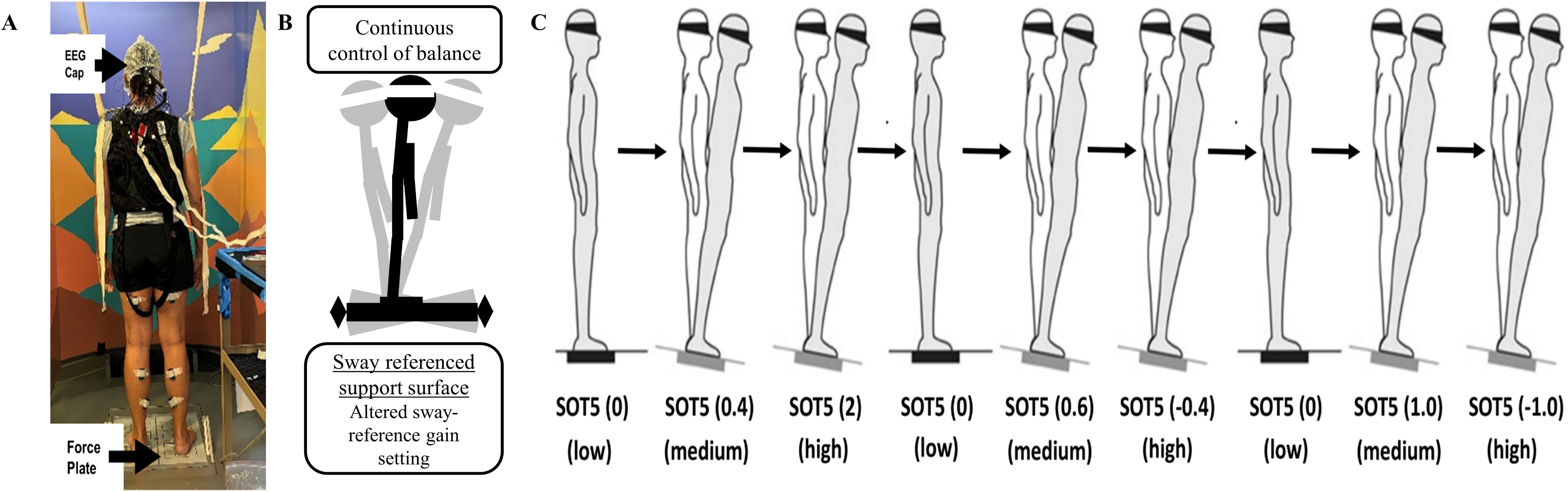
Continuous Balance Task Experimental Paradigm. Fig. 1a. Experimental setup for EEG testing during the continuous balance task. Fig. 1b. Sway references support surface. Fig. 1c. Continuous Balance task.

The continuous balance task lasted for 180 s, with three 20 s testing conditions at a gain of 0 and one 20 s condition for each of the other four gain values presented in the following order for each participant: 0, 0.6, −0.4, 0, 1.0, −1.0. During the task, participants were instructed to maintain their balance. The order of these conditions was chosen so that we first had a *low* difficulty condition with a stable support surface (gain value of 0), and then the difficulty was gradually increased to a *medium* level (gain values of 0.6 or 1.0), and then increased further to a *high* level (gain values of −0.4 or −1.0). The general pattern of *low*, *medium*, and *high* difficulty was repeated three times. These varying balance conditions allowed us to gradually increase the difficulty of the balance control task while simultaneously monitoring the EEG, EMG, and GRF responses. Thus, this task was developed with the specific intent of challenging the balance control system in such a way that both its behavioral (GRF measures) and neuromuscular (EEG and EMG measures) underpinnings could be observed during the stable (low difficulty), cautious (medium difficulty), and threatened (high difficulty) stages of balance control. Participants were not informed when the gain changed. To ensure safety, participants always wore a safety harness. In addition, they also wore a physical therapy belt with a spotter standing nearby to prevent any falls. Participants were allowed to practice at gain values of 0, 0.4, and 2.0 prior to exposure to the start of the data collection. Data from these gain values were not used for analysis.

### Electroencephalography (EEG)

64 active channel EEG electrodes (Brain Products GmbH, Germany; 1000 Hz) were used to record Whole-scalp electroencephalography (EEG). The electrodes were placed as per the International 10-20 system for EEG electrode placement. For instance, to locate Cz, four fiducial points, namely nasion, inion, left preauricular, and right preauricular, were identified. Cz was identified at the midpoint of the nasion-inion arc length and the left-right preauricular arc length. Four of the electrodes were used to record electrooculography (EOG) signals. EEG was recorded at rest and during the balance task. We constrained the movement of the EEG cables by placing an elastic mesh on the top of the EEG cap, which aided in minimizing motion artifacts during the balance task on the Neurocom [44,47]. We utilized a modified international 10-20 EEG system, where we moved the GND and REF from the default locations (AFz and FCz) to the left and right earlobes, respectively [48]. We made these modifications because the default channel locations for GND and REF are very close to the fronto-central cortex, a region of interest for this study. The empty spots were filled by moving T7 and T8 electrodes to AFz and FCz locations, respectively.

### Electromyography (EMG)

We recorded activity of thigh and lower leg muscles bilaterally using differential surface electrodes from tibialis anterior (TA), gastrocnemius medialis (GM), soleus (SOL), rectus femoris (RF), and biceps femoris (BF) (1111.11 Hz, gain 1000; 20-450 Hz Delsys Trigno EMG System, Boston, MA). These muscles were selected because of their involvement in the control of vertical posture while dealing with symmetrical perturbations induced in the sagittal plane [49,50]. The placement of electrodes for recording EMG activity was based on recommendations reported in the literature [49].

### Experimental Protocol

Each participant participated in a single session. During the session, first, we obtained their clinical measures using the BBS and TUG tests. Following this step, we prepared them for EEG and EMG measurements. Before electrode placement, the skin surface was prepared for EMG by cleaning with isopropyl alcohol pads. Each participant then performed the challenging balance task with varying sensory conditions while standing on the force platform (support surface). We obtained baseline measurements for 60 sec during quiet standing with eyes closed before and after the balance task. The dynamic balance task consisted of maintaining an upright stance with eyes closed during a nine sequential 20 sec duration balance task conditions.

### Data Analyses

#### Behavioral data

BBS (out of 56) and TUG (average of three trials in seconds) were collected before the balance task.

The ground reaction force data collected from the force plate were combined to create a center of pressure (COP) time series in medio-lateral (ML) and anterior-posterior (AP) directions for the continuous balance task [51]. Next, we estimated the COM by low-pass filtering the COP data (second-order Butterworth; f_c_ = 0.86 Hz) [44,52] for both directions of the continuous balance task. Several studies have confirmed the reliability of this method to estimate COM from COP [53–55]. We computed various linear COP measures, including mean, standard deviation, and root mean square (RMS) of COP [56] to quantify spatial measures of balance performance, as well as COP velocity measures (mean, standard deviation, and RMS of COP velocity) [57], which are also a reliable measure in the quantification of balance control. Additionally, we computed path length (PL) [58] from COP [59–61], representing cumulative COP displacement, and Sample entropy (SE) as a non-linear measure of balance control, providing a comprehensive postural performance assessment across varying task conditions.

The 95% confidence ellipse area was computed as an additional metric of balance stability, capturing the two-dimensional variability of COP trajectories in the anterior–posterior (AP) and mediolateral (ML) directions across low, medium, and high difficulty conditions. This measure reflects the spatial distribution of sway by constructing an ellipse that encloses ∼95% of the data points, offering a more comprehensive assessment than linear measures such as path length. The covariance matrix (Σ) was derived from AP and ML COP variances and their covariance, and the ellipse area was calculated as:

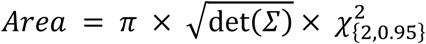

where det(Σ) is the determinant of the covariance matrix, and 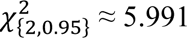 is the critical value from the chi-square distribution with 2 degrees of freedom at the 95% confidence level [62,63]. This method was applied separately for each difficulty condition, allowing for a detailed comparison of stability across low (gain = 0), medium (gains = 0.4, 0.6, 1.0), and high (gains = - 0.4, −1.0, 2.0) levels, with larger ellipse areas indicating greater sway and reduced stability[62,63].

The number of falls induced by platform translations was also recorded using qualitative observations during testing. A fall was noted when a participant lost his/her balance and required either self-induced stepping and/or support from the overhead harness to prevent them from falling to the floor (e.g., participants applied force through the rope and harness system as evident from the rope and harness going taut). A spotter was instructed to assist the participant only after they had fallen (i.e., to prevent swinging caused by the rope).

#### EEG pre-processing

The EEG pre-processing pipeline used in this study was similar to that described in our previous studies [13,44,64,65]. The raw EEG signals sampled at 1000 Hz were first downsampled to 250 Hz. EOG channels (4 channels) were utilized for adaptive filtering of the EEG signals through the H infinity algorithm with gamma parameter = 1.1 and q parameter set as 1e-11 to remove eye artifacts, correct baseline drifts, and remove other shared sources of noise [66]. A Zapline function in EEGLab was used to remove the line noise[67]. A standardized early-stage EEG processing pipeline (PREP) with default parameters was used to remove artifactual EEG channels and apply a robust common referencing method to increase the signal-to-noise ratio [68]. PREP is reported to be more robust in detecting noisy channels compared to the previously known noisy channel detection method: *pop_rejchan* from EEGLAB [68,69]. The PREP pipeline also replaces artifactual channels with surrogate data that is interpolated from neighboring sensors. This minimizes bias when performing common average referencing. The signals were then band-pass filtered at 0.1-100 Hz using a 4th-order Butterworth filter to remove slow-drifting noise. Artifact Subspace Reconstruction (ASR) was applied next to detect and denoise any artifactual sections in the EEG data [70]. The ASR uses clean EEG data to calibrate noise covariance and uses a standard deviation threshold of 15 with a sliding window (500 ms) based principal component analysis (PCA) to detect noisy sections in the EEG data [70–72]

Next, to compensate for the rank deficiency in the data, surrogate channels during the PREP pipeline were removed before running the independent component analyses (ICA), since the ICA assumes the number of independent components to be the same as the number of channels provided as the data. Adaptive mixture ICA (AMICA) was used to compute the maximally independent components (ICs) from the data [73]. The AMICA is reported to be the best ICA method available to date, and it provides more dipolar and minimized mutual information ICs [74,75]. Using the constructed boundary elemental model (BEM) explained in the next paragraph and the digitized channel locations, the dipole fitting method: *DIPFIT* in EEGLAB [69,76] was used to calculate the equivalent current dipole sources that explain at least 85% [77] of topographic variance obtained from ICA results. The digitized channel locations were first warped onto the constructed head model before calculating the dipole locations. We removed dipoles located outside the individual BEM model, as well as those with artifactual components such as muscle-related power spectral density characteristics and motion artifact-related high-frequency noise.

The boundary element model (BEM) was created after computing surface meshes and constructing the head model using a standard MRI template. The constructed BEM was then saved and used as one of the arguments in the *DIPFIT* process. Each IC scalp projection, its equivalent dipole’s location, and its power spectra were then visually inspected, and ICs that related to non-brain artifacts (e.g., sensor movement, muscle artifact, stimulation artifact) were removed.

#### EMG pre-processing

The electromyography (EMG) signals were preprocessed to ensure high-quality data for analysis. Raw EMG data, recorded bilaterally from tibialis anterior (TA), gastrocnemius medialis (GM), soleus (SOL), rectus femoris (RF), and biceps femoris (BF) at a sampling rate of 1111.11 Hz, were downsampled to 250 Hz to align with the EEG sampling rate. The signals were detrended to remove linear trends and filtered using a built-in bandpass filter (20-450 Hz) to isolate muscle activation frequencies while reducing noise. Artifactual segments, such as those corrupted in the final two seconds of several trial conditions, were excluded from further analysis. This preprocessing ensured that only reliable EMG data were used for subsequent wavelet coherence analysis.

### Wavelet coherence analysis

Wavelet coherence (WC) was used to investigate the relation in time-frequency space between EEG and EMG signals. It is a method of measuring the cross-correlation between two signals with respect to time and frequency. We computed the WC between the Cz (EEG) electrode and EMG activity measured from TA, GM, SOL, BF, and RF using signals from both sides in all the participants. We selected Cz electrode location because it is known to represent the sensorimotor area of the cortex overlying the leg representation [18,78,79]. We used the Morlet wavelet as the mother wavelet, with default parameters set as one octave for 10 scales, as it provides good time and frequency localization, and a complex shape that allows for capturing both amplitude and phase information [80]. The localized coherence significance was evaluated using Monte Carlo analysis with 1000 iterations, as this provides a way to estimate statistical significance and assess the reliability of the wavelet results. WC was computed over the first 18 sec of trial (4500 samples) with a frequency resolution of 0.0526 Hz. To understand the dynamic nature of coherence between EEG and EMG, WC was averaged over a 2-second nonoverlapping trial segment (9 bins). Values outside the cone of influence (COI) were considered highly unreliable and thus excluded from the analysis [80]. This also led to the exclusion of the first and last bins. Coherence was considered significant (p < 0.05) if it was greater than Z, which is given using the following formula:

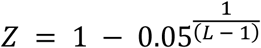

where L is the total number of non-overlapping segments, equal to the available number of trials (6 trials; practice trials not included) multiplied by seven [81], resulting in a Z value of 0.07. The average coherence values were then converted to Fisher z-statistics (normalized) using the arc hyperbolic tangent transformation [82,83] prior to performing any statistical analysis. We analyzed corticomuscular coherence in delta (0.05 to 4 Hz), theta (4 to 8), alpha (8-12 Hz), beta (12-30 Hz), and gamma (30-50 Hz) frequency bands. Although the EMG signals were filtered with a built-in bandpass filter, it is not unusual to experimentally observe significant coherence at lower frequencies [13,84–86]

### Composite Variables

a) <u>Asymmetry Index (AI)</u> The corticomuscular coherence (CMC) asymmetry index was calculated to quantify motor control imbalance between the affected side and the less affected side in stroke patients and between the dominant and non-dominant side in healthy participants. CMC values were derived using wavelet coherence between the EEG at Cz and EMG signals from corresponding muscles. For stroke participants, the AI was computed as the ratio of coherence from the affected vs. unaffected side, while in healthy participants, it was calculated for the dominant vs. non-dominant side. The CMC asymmetry index (AI) for both groups was determined using the formula:

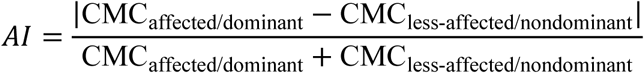

where CMC_affected/dominant_ represents the average coherence values for the affected or dominant side and the CMC_less-affected/nondominant_ represents the average coherence values for the unaffected or non-dominant side, respectively, across specified frequency bands (e.g., delta to gamma). The resulting index ranges from 0 to 1, where 0 indicates perfect symmetry in coherence between sides, and 1 signifies complete asymmetry, reflecting potential motor control differences. Elevated AI reflects corticospinal disruption in stroke, while smaller asymmetries in healthy individuals capture natural lateralization. Prior studies have demonstrated increased CMC asymmetry in stroke participants, associated with motor deficits [87–89], whereas minor asymmetries in healthy participants may be attributed to leg dominance [90].

b). <u>Task Modulation Index (TMI):</u> The corticomuscular coherence (CMC) task modulation index (TMI) was computed to evaluate motor system adaptability across varying balance task difficulties (low, medium, high). CMC was estimated via wavelet coherence between EEG at Cz and EMG from lower-limb muscles (tibialis anterior, gastrocnemius medialis, soleus, rectus femoris, biceps femoris). TMI was derived for two contrasts (high–medium, medium–low) using the standard formula:

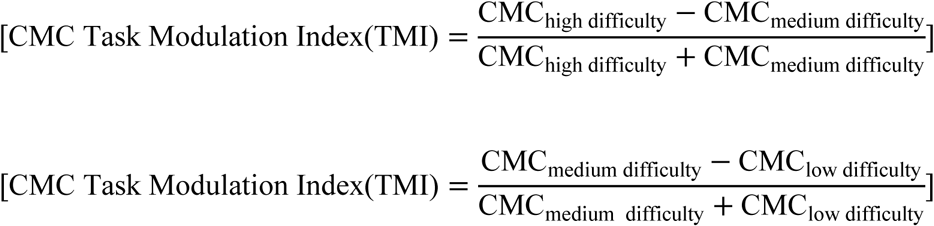

where CMC_high difficulty_, CMC_medium difficulty_, and CMC_low difficulty_ represent the average coherence values for the high (gains = −1.0, 2.0), medium (gains = 0.6, 1.0), and low (gain = 0) difficulty conditions, respectively, across relevant/all frequency bands. The TMI values range from −1 to 1, with positive values indicating an increase in coherence with higher difficulty, negative values suggesting a decrease, and values near zero reflecting minimal change, thus capturing the relative modulation of coherence across task demands. This measure reflects the capacity of cortical motor networks to adapt under increasing task demands and is clinically relevant for stroke rehabilitation. Prior studies confirm that task-dependent modulation of CMC reflects corticomotor connectivity changes and adaptive control [13,91] with implications for motor recovery.

c). <u>Area under Curve (AUC):</u> The area under the coherence curve (AUC) was calculated within standard frequency bands (delta 0.1–4 Hz, theta 4–8 Hz, alpha 8–12 Hz, beta 12–30 Hz, gamma 30–50 Hz) to quantify the overall strength of corticomuscular interaction. AUC was obtained by integrating the wavelet coherence spectrum (EEG Cz–EMG pairs) using the trapezoidal rule applied to the coherence spectrum obtained from the Cz electrode EEG and EMG signals (e.g., tibialis anterior, gastrocnemius medialis, soleus, rectus femoris, and biceps femoris). The AUC was computed as:

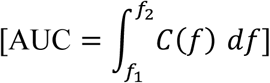

where C(f) represents the coherence function over the frequency range from f_1_ to f_2_ (the boundaries of each frequency band), approximated numerically using the trapezoidal method based on the sampled coherence data. Unlike traditional CMC analyses that focus on peak or mean coherence at a specific frequency, which captures the strength of coupling at one point in the spectrum, CMC AUC integrates coherence across the entire frequency band, capturing both the magnitude and extent of corticomuscular interaction. This cumulative approach provides a robust estimate of neuromuscular coupling and is particularly useful for relative contributions of each frequency band to balance control. Prior work has emphasized the role of frequency-specific corticomuscular coherence in rhythmic motor communication [92] and the statistical rigor of coherence-based spectral analyses [93]; however, integrating AUC provides a more holistic view of corticomuscular interactions, offering insights into overall coherence across frequency bands rather than focusing on individual frequencies. Additionally, stroke patients do exhibit altered frequency profiles compared to healthy controls [13,94], suggesting that CMC AUC can serve as a sensitive biomarker in distinguishing normal and pathological neural strategies.

### Machine Learning Analysis: Data Preparation

The machine learning analysis utilized a dataset comprised of neurophysiological and biomechanical features derived from EEG, EMG, and COP measurements during balance tasks. Extracted variables included corticomuscular coherence (CMC) asymmetry indices, task modulation indices, area under the coherence curve (delta–gamma bands), COP 95% confidence ellipse area, Berg Balance Scale (BBS), Timed Up and Go (TUG), age, and subject identifiers. The cohort included 33 participants (18 stroke patients, 15 healthy controls).

Missing values, resulting from falls or assistance during trials, were identified using a comprehensive assessment of numerical features. To address these, the dataset was stratified into stroke and healthy subgroups based on subject identifiers, and missing data were imputed using the k-nearest neighbors (KNN) imputation method, which estimates missing values based on the similarity of feature vectors within each group [95]. Imputation was performed separately for each subgroup to preserve group-specific patterns. A grid search over n_neighbors values (1, 3, 5, 7, 9) was conducted, with n=9 selected as optimal, as it minimized the number of columns with imputed values significantly deviating from original group means (p < 0.05, evaluated using one-sample t-tests) while maximizing n_neighbors to reduce imputation variance.

A binary ‘Group’ column was introduced, assigning 0 to stroke participants and 1 to healthy controls, to facilitate classification tasks. The preprocessed dataset, validated for the absence of missing values and comprising 33 rows and 528 columns (**Table 2**)—including age, behavioral measures, and EEG-EMG coherence variables across five muscles per side (biceps femoris, rectus femoris, tibialis anterior, gastrocnemius medialis, soleus) under low, medium, and high task conditions—was exported as a new Excel file. This approach ensured robust data handling for subsequent analyses, adhering to established guidelines for managing missing data in biomedical research [96].

**Table 2.** Features used in the study. Muscle abbreviations: BF - Biceps Femoris; RF - Rectus Femoris; TA - Tibialis Anterior; GM - Gastrocnemius Medialis; SOL - Soleus.

| Feature | Conditions | Side | Muscles | Frequency band | Count |
| --- | --- | --- | --- | --- | --- |
| Cortico-muscular Coherence (CMC) | Low (lowcomb), Medium (medcomb), High (highcomb) | LN-Less affected stroke (L) paired with non-dominant (N) of healthy.<br>AD/AR-Affected stroke (A) paired with dominant (D) of healthy. | BF, RF, TA, GM, SOL | Delta (D), theta (T), alpha (A), beta (B), gamma (G) | 150 |
| CMC Asymmetry Index (AI) | Low (lowcomb), Medium (medcomb), High (highcomb) | N/A | BF, RF, TA, GM, SOL | Delta (D), theta (T), alpha (A), beta (B), gamma (G) | 75 |
| CMC Task Modulation Index (TMI) | High→Medium (HM), Medium→Low (ML) | LN, AD | BF, RF, TA, GM, SOL | Delta (D), theta (T), alpha (A), beta (B), gamma (G) | 100 |
| Cortico-muscular Coherence Area Under Curve (AUC) | Low (lowcomb), Medium (medcomb), High (highcomb) | LN, AD | BF, RF, TA, GM, SOL | Delta (D), theta (T), alpha (A), beta (B), gamma (G) | 150 |
| COP Features – Anterior-Posterior (AP) direction (Mean, Std, RMS, Velocity, Entropy) | Low (lowcomb), Medium (medcomb), High (highcomb) | N/A | N/A | N/A | 15 |
| COP Features – Medio-Lateral (ML) direction (Mean, Std, RMS, Velocity, Entropy) | Low (lowcomb), Medium (medcomb), High (highcomb) | N/A | N/A | N/A | 15 |
| COP Path length (COP_PL) | Low (lowcomb), Medium (medcomb), High (highcomb) | N/A | N/A | N/A | 15 |
| COP 95% Confidence Ellipse Area (EllipseArea) | Low (lowcomb), Medium (medcomb), High (highcomb) | N/A | N/A | N/A | 3 |
| Subject | N/A | N/A | N/A | N/A | 1 |
| Group | N/A | N/A | N/A | N/A | 1 |
| Age | N/A | N/A | N/A | N/A | 1 |
| Berg Balance Score (BBS) | N/A | N/A | N/A | N/A | 1 |
| Timed-Up-And-Go (TUG) | N/A | N/A | N/A | N/A | 1 |
|  |  |  |  |  | **Total = 528** |

### Feature Selection (via Classification) for Regression

The dataset presented a high-dimensional, small-sample challenge (528 features, n = 33; **Table 2**), where direct regression to predict BBS and TUG scores would risk severe overfitting and poor generalizability. To address this, we adopted a two-stage pipeline. In the first stage, binary classification models (stroke versus healthy) were applied to perform feature selection. This step will identify variables that best discriminate between groups (Stroke versus Healthy). Classification-based feature selection has been widely recommended in machine learning to reduce dimensionality while preserving interpretability [35,36]. In the second stage, the selected features will be entered into regression models to predict continuous BBS and TUG scores, ensuring that the subsequent analyses are based on physiologically meaningful and statistically robust variables. This pipeline leverages classification not only to identify biomarkers that separate stroke from healthy controls but also to improve the stability and interpretability of regression models aimed at functional outcome prediction.

### Machine Learning Analysis (Classification Task)

The classification task aimed to identify clinically significant features for differentiating stroke patients from healthy controls, serving as a feature selection mechanism to guide subsequent regression analysis for predicting Berg Balance Scale (BBS) and Timed Up and Go (TUG) scores, by conducting a thorough evaluation of model performance across a diverse set of algorithms. Seven machine learning models were selected to represent key algorithmic categories: Ensemble Methods (XGBoost, Random Forest, AdaBoost) for their robustness to noise, ability to handle non-linear relationships, and feature importance [97]; Support Vector Machine (SVM) for its effectiveness in high-dimensional spaces via margin maximization and kernel tricks, and Regression Methods (Logistic Regression with L1, L2, and Elastic Net regularization) for their interpretability, sparsity induction and regularization to prevent overfitting in linear models [98]. This diverse ensemble ensured a comprehensive assessment of predictive performance, facilitating the identification of optimal models without category bias. Hyperparameter optimization using GridSearchCV with 3-fold stratified cross-validation (StratifiedKFold (n_splits=3, shuffle=True, random_state=42)), enhanced parameter tuning robustness while preserving class distribution [99]. Parameter grids were customized to each model’s complexity, such as max_depth (2-4) and learning_rate (0.01-0.2) for XGBoost, n_estimators (50-200) and max_depth (2-10) for Random Forest, and C (0.001-10) with kernel options (linear, rbf) for SVM [100]. To reduce variability from stochastic processes and ensure stability and reproducibility, models were trained and evaluated across three distinct random seeds (42, 100, 123), to produce reliable performance assessment in machine learning [98,101].

The preprocessed dataset was partitioned into training (24 samples) and testing (9 samples) sets using a stratified 75-25 split with random_state=42 to maintain the original class ratio[102]. Features were standardized using StandardScaler for compatibility with scale-sensitive models like SVM and Logistic Regression. Performance was evaluated using accuracy, precision, recall, F1 score, and ROC AUC, alongside confusion matrices for detailed insights across all model-seed combinations [103]. SHAP values were computed for interpretability—using TreeExplainer for ensembles, KernelExplainer for SVM, and LinearExplainer for Logistic Regression—extracting features with non-zero coefficients from both training and test sets [104]. Model performance metrics were aggregated into a DataFrame, calculating means and standard deviations for key indicators (Test ROC AUC, Test F1, Test Recall, Test Accuracy) across seeds, with robust error handling [100]. The top two models were selected based on the highest average Test ROC AUC, using standard deviation for consistency, supported by visualizations to prioritize overall predictive power and stability for subsequent feature analysis [98,101].

### Feature Importance Consolidation, Selection, and UMAP

To identify key features for distinguishing stroke patients from healthy controls, feature importance was evaluated for the top two models. The preprocessed dataset was validated to ensure no missing values remained after KNN imputation, with non-numeric columns excluded using scikit-learn tools[100]. SHapley Additive exPlanations (SHAP) values, computed during the initial classification task across three random seeds (42, 100, 123), were aggregated to capture features with non-zero absolute coefficients for each model-seed combination, accounting for stochastic variability [104]. Features were ranked based on their frequency of non-zero SHAP values across models, and only features that appeared in at least four out of six model runs (i.e., more than 50% of the time) were retained, forming a standardized feature set for subsequent regression analyses. To assess how well these selected features differentiate the two groups, Uniform Manifold Approximation and Projection (UMAP) was applied to reduce the feature space to two dimensions, enabling visualization of cluster separation between stroke patients and healthy controls via scatter plots colored by group label [105]. All results were saved for feature evaluation and visualization in machine learning [98,100]

### Machine Learning Analysis (Regression Task)

Feature set identified from the classification task, along with two best-performing classification models, were used for regression to ensure consistent feature importance and reliable prediction of BBS and TUG scores. The dataset, comprising 33 subjects (18 stroke, 15 healthy) and selected features, was partitioned into training (24 samples) and testing (9 samples) sets using a stratified 75-25 split with random_state=42 to maintain group balance [102]. Features were standardized using StandardScaler for compatibility with the models. Hyperparameter tuning was performed using GridSearchCV with 5-fold cross-validation (KFold, random_state=42) to optimize model performance. Models were trained and evaluated using a single random seed (42) for reproducibility, reducing computational complexity compared to the multi-seed classification approach. Performance was assessed using R², adjusted R², root mean squared error (RMSE), mean absolute error (MAE), and F-statistic with p-values to evaluate predictive accuracy and model fit. Group-wise metrics (stroke versus healthy) were computed to assess performance differences. Residual analysis was done as well and SHAP values were calculated to quantify feature contributions [104]. Performance metrics were aggregated into Data Frames and visualizations, which were saved for further analysis.

### Feature Specific Analysis

To evaluate the contribution of the selected features predicting the Berg Balance Scale (BBS) and Timed Up and Go (TUG), feature-specific analysis was conducted using regression models (seed 42) on training (24 samples) and testing (9 samples) sets with standardized features. Computed model-specific SHAP values for the train data were utilized to quantify each feature’s impact on predictions, with features sorted by absolute mean SHAP values, where higher values indicate greater influence on model outputs[104]. Beeswarm plots were generated for train and test sets to visualize SHAP values, where wider spreads denote greater feature importance, and color/position relative to zero indicates whether higher or lower feature values increase or decrease predicted BBS/TUG scores. Spearman rank correlations were calculated between each feature and predicted BBS/TUG scores to assess monotonic relationships, with Bonferroni-adjusted p-values to account for multiple comparisons and determine statistical significance (p < 0.05).

## Results

### Data Preparation Outcomes

The dataset comprised 33 subjects (18 stroke participants and 15 healthy controls) with 528 features (**Table 2**). Missing values were handled using k-nearest neighbors (KNN) imputation with an optimized n_neighbors=9, resulting in 23 columns with imputed values significantly differing from group means (p < 0.05, one-sample t-tests). Post-imputation, the dataset was confirmed to have no missing values, ensuring readiness for subsequent classification analyses.

### Performance of the Machine Learning Models for Classification

**Table 3**, **Fig. 2a & b** summarizes the performance of seven classification models—XGBoost, Random Forest, AdaBoost, SVM, and Logistic Regression with L1, L2, and Elastic Net regularization—evaluated across three random seeds to distinguish stroke participants from healthy. Among these, Logistic Regression (Elastic Net) and XGBoost were most effective, with strongest overall performance metrics. The other models showed moderate effectiveness. Logistic Regression with L1 and L2 regularization, and Random Forest performed reasonably well, though AdaBoost lagged behind and SVM struggled the most, with poor results and high variability (**Table 3**; **Fig. 2a)**. Based on these performance metrics, Logistic Regression (Elastic Net) and XGBoost were selected for further analysis. Examining confusion matrices across the three seeds, both models performed near-perfect classification. XGBoost (Seed 42) demonstrated perfect classification, both in training and test sets (**Fig. 2b**). On the other hand, Logistic Regression (Elastic Net) (Seed 42) performed perfectly in training, with a minor misclassification in testing (**Fig. 2c**).

**Fig 2.**
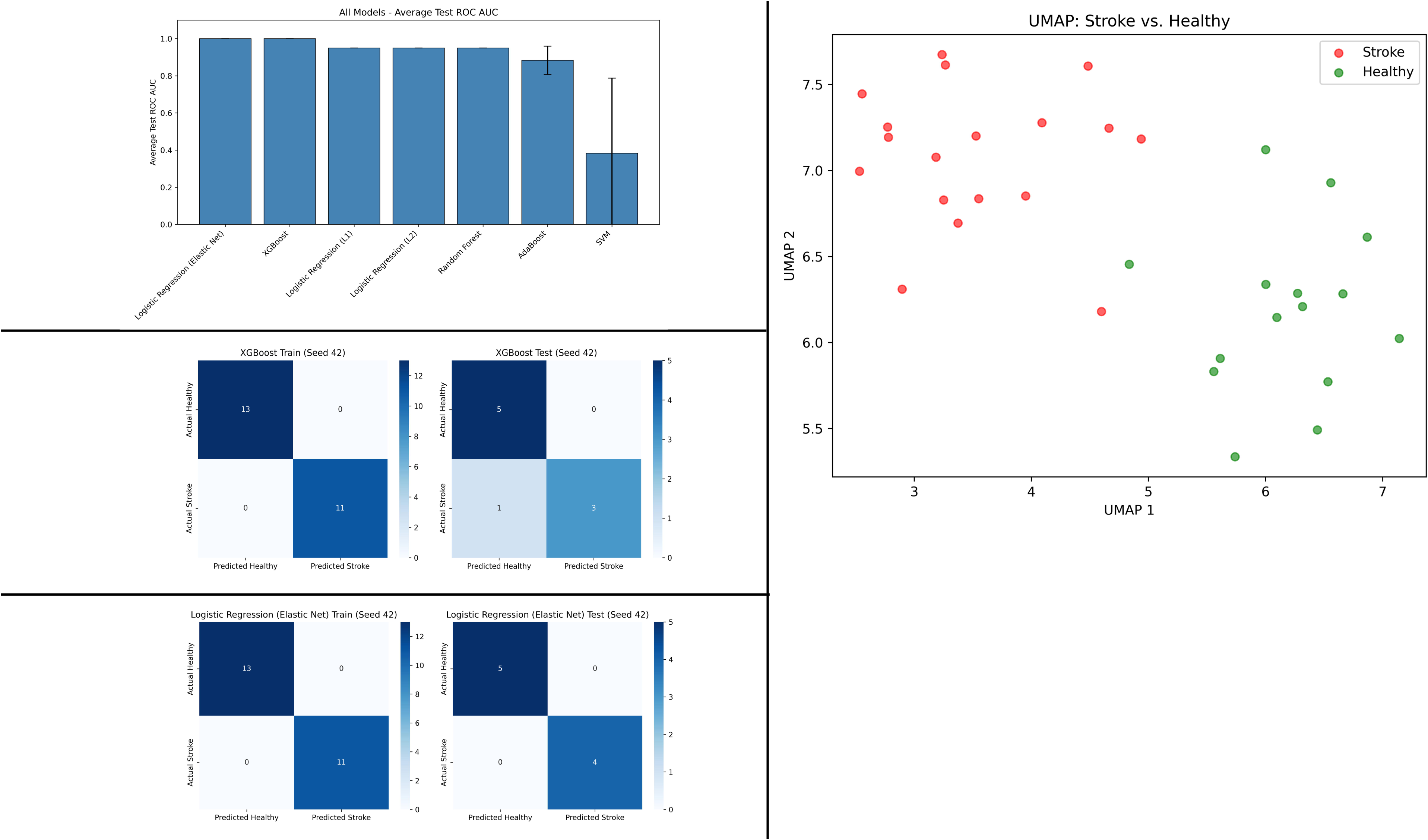
Comparative Model Performance Metrics and Dimensionality Reduction (UMAP) Analysis. Fig. 2a **(top-left panel):** Average Test ROC AUC scores (± standard deviation) for seven classification models. Fig. 2b **(middle-left panel):** Confusion matrices for XGBoost model in classifying stroke vs. healthy participants, showing training and testing results (Seed 42). Fig. 2c **(bottom-left panel):** Confusion matrices for Logistic Regression (Elastic Net) model in classifying stroke vs. healthy participants, showing training and testing results (Seed 42). Fig. 2d **(top-right panel):** UMAP projection of the dataset, with separation between Stroke and Healthy participants.

**Table 3:**
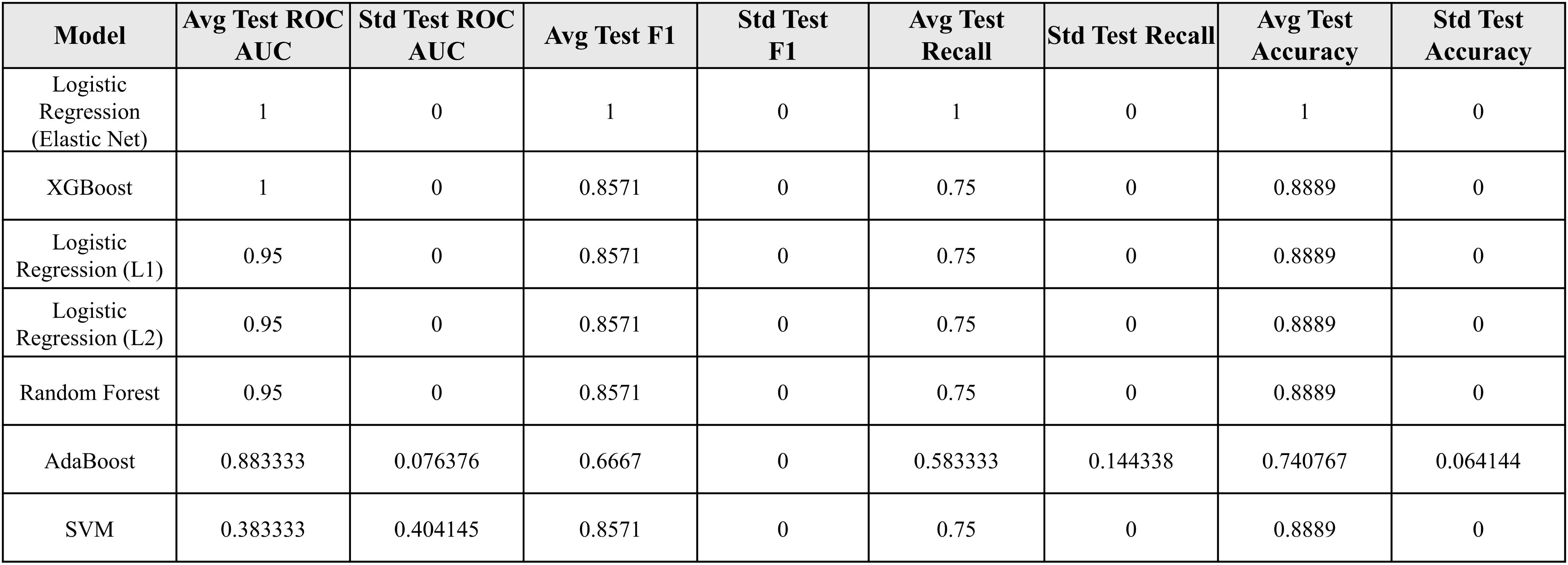
Comparative Model Performance Metrics. Aggregated Model Performance Metrics: Average and standard deviation of ROC AUC, F1 score, recall, and accuracy for test dataset across seven classification models.

### Feature Selection

To identify most important features (with non-zero SHAP coefficients), we consolidated feature importance across the top two best-performing models — evaluated over three random seeds (42, 100, 123). This produced a comprehensive list of 89 unique features. Then we applied the selection criterion: features were ranked based on the frequency of non-zero SHAP values across all model runs, and only those appearing in at least four out of six runs (i.e., more than half the time) were retained. This resulted in 15 key features, as reported in **Table 4**. These features, including CMC metrics (e.g., task modulation and asymmetry indices, AUC), balance measures (BBS, TUG), and center of pressure variables (e.g., variability measures), consistently distinguished stroke patients from healthy controls. Dimensionality reduction via Uniform Manifold Approximation and Projection (UMAP) on this refined set revealed clear cluster separation (**Fig. 2d)**, with stroke subjects (red) predominantly on the left and healthy subjects (green) on the right, underscoring the discriminative power of the selected features.

**Table 4:** Feature Selection. Cumulative frequency of feature selection across Logistic Regression (Elastic Net) and XGBoost models.

| Feature | Logistic Regression frequency | Xgboost frequency | Total frequency |
| --- | --- | --- | --- |
| RF CMC for less affected Stroke side with non-dominant Healthy side for beta band in high condition | 3 | 3 | 6 |
| Timed-Up-and-Go | 3 | 3 | 6 |
| TMI of GM CMC for Affected Stroke side with dominant Healthy side for beta band from medium to low condition | 3 | 2 | 5 |
| AI of SOL CMC for theta band in medium condition | 3 | 2 | 5 |
| AI of BF CMC for beta band in high condition | 3 | 2 | 5 |
| TMI of RF CMC for Affected Stroke side with dominant Healthy side for beta band from high to medium condition | 3 | 2 | 5 |
| AI of RF CMC for gamma band in high condition | 3 | 2 | 5 |
| Standard deviation of COP Standard deviation in Medio-lateral direction for high condition | 3 | 2 | 5 |
| Berg Balance Score | 2 | 3 | 5 |
| AI of RF CMC for beta band in high condition | 3 | 1 | 4 |
| Sample Entropy of COP in Medio-lateral direction-for high condition | 3 | 1 | 4 |
| AI of TA CMC for theta band in medium condition | 3 | 1 | 4 |
| TMI of TA CMC for Affected Stroke side with dominant Healthy side for delta band from high to medium condition | 3 | 1 | 4 |
| Root mean square of COP in Medio-lateral direction for high condition | 3 | 1 | 4 |
| AUC of TA CMC for Affected Stroke side with dominant Healthy side for delta band for medium condition | 3 | 1 | 4 |

### Regression results

The classification task served as a feature selection mechanism to guide subsequent regression modeling for predicting Berg Balance Scale (BBS) and Timed Up and Go (TUG) scores. The identified 15 features included the BBS and Timed Up and Go TUG. So, for the regression task, the remaining 13 features served as predictors in regression models to estimate BBS and TUG scores. XGBoost and Logistic Regression with Elastic Net regularization, selected for their best performance in classification, were employed to ensure consistency in feature importance and robust predictive performance for these functional outcomes.

### Regression Model Performance

The performance of XGBoost and Elastic Net regression models, using 13 selected features to predict Berg Balance Scale (BBS) and Timed Up and Go (TUG) scores, was evaluated on training (24 samples) and testing (9 samples) sets (**Fig 3**). For BBS, the XGBoost model achieved near-perfect training performance (RMSE = 0.0358, **Fig 3a**) but lower test performance (RMSE = 5.0520, **Fig 3b**). For BBS, the Elastic Net model showed moderate training (RMSE = 4.2511, **Fig 3a**) but poor test generalization (RMSE = 5.6372, **Fig 3b**).

**Fig 3.**
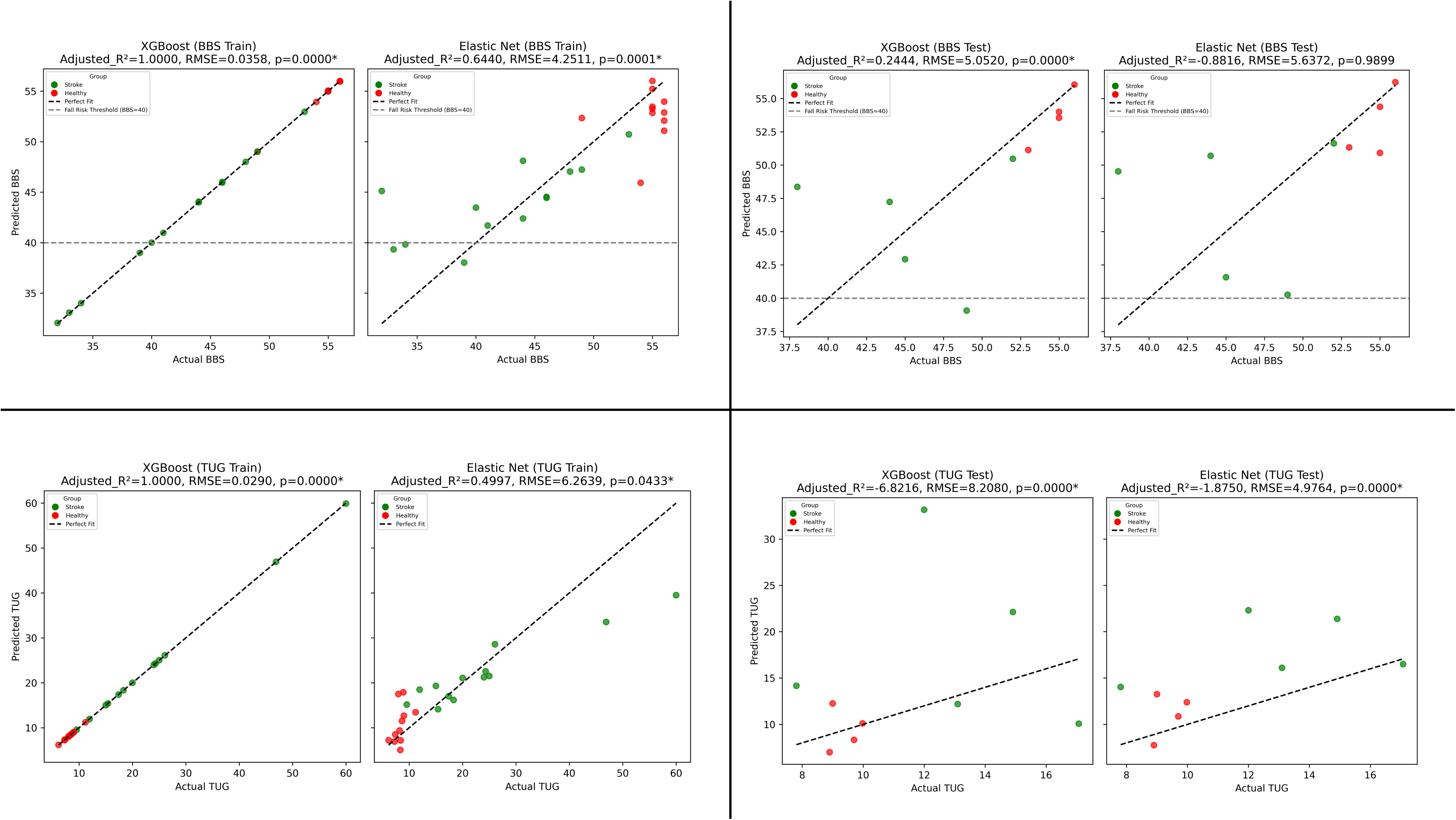
BBS and TUG Prediction Model Performance. Fig. 3a **(top-left panel):** XGBoost & Elastic Net BBS prediction performance on the training dataset. Fig. 3b **(top-right panel):** XGBoost & Elastic Net BBS prediction performance on the test dataset. Fig. 3c **(bottom-left panel):** XGBoost & Elastic Net TUG prediction performance on the training dataset. Fig. 3d **(bottom-right panel):** XGBoost & Elastic Net TUG prediction performance on the test dataset. Points are colored by group: “Stroke” (red) and “Healthy” (green).

For TUG, the XGBoost model exhibited excellent training fit (RMSE = 0.0290, **Fig 3c**) but poor test performance (RMSE = 8.2080, **Fig 3d**). For TUG, the Elastic Net model showed moderate training performance (RMSE = 6.2639, **Fig 3c**) but negative test generalization (RMSE = 4.9764, **Fig 3d**). Negative adjusted R² values on the test set indicate models performed worse than a mean-based baseline, likely due to overfitting and small test sample size (n=9).

### Feature Selection for the Prediction of BBS

For BBS prediction using the XGBoost model, we rank-ordered all 13 features based on their absolute SHAP coefficients (**Fig 4a)** and then examined the correlation of each feature with BBS scores, applying Bonferroni correction to adjust for multiple comparisons. Three features exhibited statistically significant Spearman correlations (p < 0.05): AUC of TA delta-band CMC on the affected stroke-dominant healthy side for the medium condition (absolute mean SHAP = 2.2857, ρ = 0.6330, p = 0.0117), asymmetry index of SOL theta-band CMC for the medium condition (SHAP = 2.1715, ρ = −0.5930, p = 0.0293), and RMS of COP in the medio-lateral direction for the high condition (SHAP = 1.6505, ρ = −0.6993, p = 0.0019) (**Figs 4a and 4c)**. The beeswarm plot (**Fig 4b**) demonstrates the influence of individual feature values on BBS predictions. Specifically, higher feature value AUC of TA CMC in delta band push predictions towards higher BBS scores, indicating that higher feature value is associated with better balance performance. In contrast, higher feature value for the asymmetry index for SOL CMC in the theta band for medium condition pushes predictions towards lower BBS scores, suggesting greater asymmetry leads to poor balance. Similarly, larger RMS of COP in the medio-lateral direction in high condition also drives towards lower BBS scores, demonstrating that increased sway in that direction reduces balance ability. showing negative correlations with BBS. Correlation plots **(Fig 4c)** further validate these directional relationships, highlighting a strong influence of these features on the prediction of BBS.

**Fig 4.**
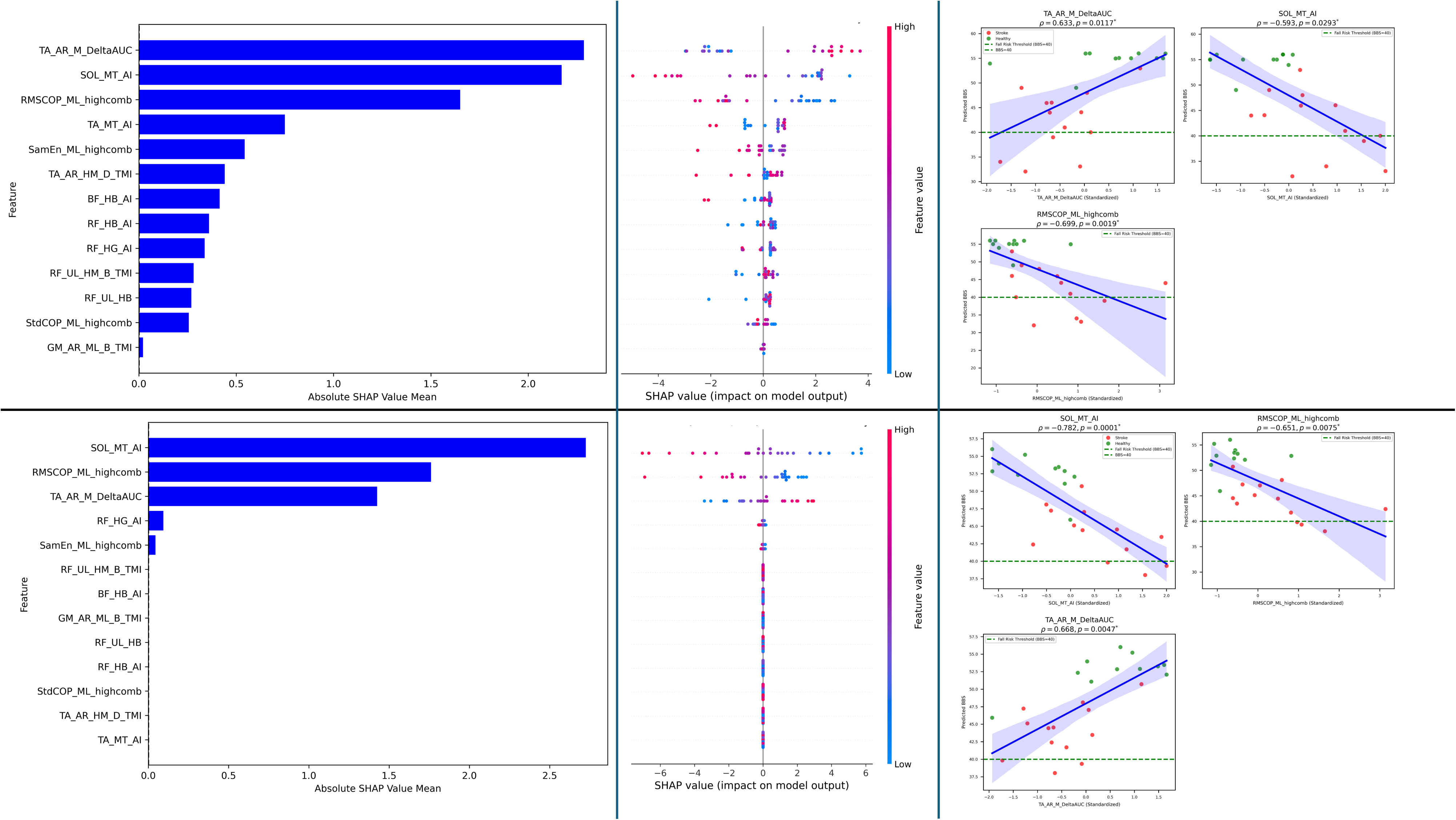
Feature analysis for BBS Prediction. Fig. 4a **(top-left panel):** XGBoost feature importance ranked by mean absolute SHAP values. Fig. 4b **(top-middle panel):** SHAP beeswarm plot showing feature impacts on XGBoost predictions (color = feature value). Fig. 4c **(top-right panel):** Correlation between standardized features and XGBoost-predicted BBS (for statistically significant features), colored by group: Stroke (red) and Healthy (green). Fig. 4d–f **(bottom panel):** Same as in Fig. 4a–c, for the Logistic Regression (Elastic Net) model.

Similar correlation findings were observed with the Elastic Net model for the same three features (p < 0.05): Asymmetry index of SOL theta-band CMC for the medium condition (SHAP = 2.2749, ρ = −0.7817, p = 0.0001), RMS of COP in the medio-lateral direction for the high condition (SHAP = 1.7611, ρ = −0.6506, p = 0.0075), and AUC of TA delta-band CMC on the affected stroke side-dominant healthy side for the medium condition (SHAP = 1.4251, ρ = 0.6678, p = 0.0047) (**Figs 4d and 4f)**. The beeswarm plot (**Fig 4e**) confirms the importance of these features with wide spreads, showing similar prediction patterns with those observed for the XGBoost model. Corresponding correlation plots (**Fig 4f)** validate these findings, demonstrating similar directional relationships highlighting the strong influence of these features on BBS prediction.

### Feature Selection for the prediction of TUG

For TUG prediction using the XGBoost model, we followed the similar process as for BBS: rank-ordered all features based on their absolute SHAP coefficients (**Fig 5a**) and then examined the correlation of each feature with TUG scores, applying Bonferroni correction to adjust for multiple comparisons. Five features exhibited statistically significant Spearman correlations (p < 0.05): asymmetry index of BF beta-band CMC in the high condition (absolute mean SHAP = 3.4580, ρ = 0.6183, p = 0.0167), RMS of COP in the medio-lateral direction for the high condition (SHAP = 3.3701, ρ = 0.6780, p = 0.0035), AUC of TA delta-band CMC on the affected stroke-dominant healthy side for the medium condition (SHAP = 2.6861, ρ = −0.6817, p = 0.0032), task modulation index of RF beta-band CMC for the less-affected stroke—non-dominant healthy side from high to medium condition (SHAP = 1.9034, ρ = −0.5974, p = 0.0267), and RF beta-band CMC for the less-affected stroke—non-dominant healthy side for the high condition (SHAP = 0.4674, ρ = −0.5974, p = 0.0267) (**Figs 5a and 5c**). The beeswarm plot (**Fig 5b**) demonstrates the influence of individual feature values on TUG predictions. Specifically, higher values of the asymmetry index for BF beta-band CMC and RMS of COP in the medio-lateral direction push predictions toward higher TUG scores, indicating that greater asymmetry and increased sway are associated with poorer mobility performance. In contrast, higher values for AUC of TA delta-band CMC, RF beta-band CMC, and task modulation index of RF beta-band CMC itself push predictions toward lower TUG scores, reflecting that stronger coherence and its task-related modulation for these features linked to better mobility. Correlation plots (**Fig 5c**) further validate these directional relationships, highlighting the strong influence of these features on TUG prediction.

**Fig 5.**
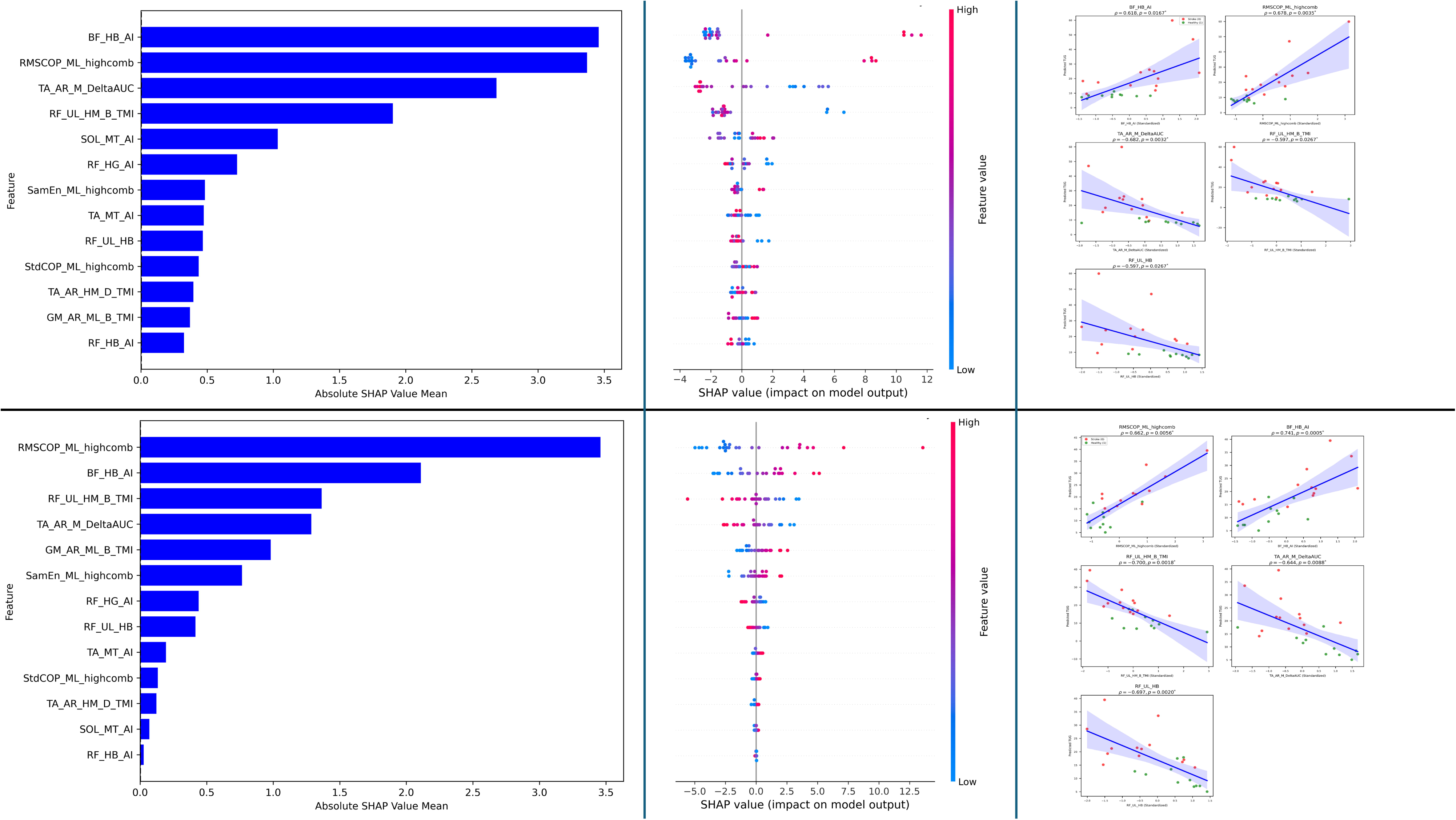
Feature analysis for TUG Prediction. Fig. 5a **(top-left panel):** XGBoost feature importance ranked by mean absolute SHAP values. Fig. 5b **(top-middle panel):** SHAP beeswarm plot showing feature impacts on XGBoost predictions (color = feature value). Fig. 5c **(top-right panel):** Correlation between standardized features and XGBoost-predicted TUG (for statistically significant features), colored by group: Stroke (red) and Healthy (green). **Fig. 5d–f (bottom panel):** Same as in Fig. 5a–c, for the Logistic Regression (Elastic Net) model.

Similar correlation findings were observed with the Elastic Net model for the same five features (p < 0.05): RMS of COP in the medio-lateral direction for the high condition (SHAP = 3.4584, ρ = 0.662, p = 0.0056), asymmetry index of BF beta-band CMC in the high condition (SHAP = 2.1097, ρ = 0.74, p = 0.00045), task modulation index of RF beta-band CMC for the less-affected stroke—non-dominant healthy side from high to medium condition (SHAP = 1.3640, ρ = −0.70, p = 0.0018), AUC of TA delta-band CMC on the affected stroke-dominant healthy side for the medium condition (SHAP = 1.2866, ρ = −0.644, p = 0.0088), and RF beta-band CMC for the less-affected stroke—non-dominant healthy side for the high condition (SHAP = 0.4160, ρ = −0.697, p = 0.0019) (**Figs 5d and 5e**). The beeswarm plot (**Fig 5d**) confirms the importance of these features with wide spreads, showing prediction patterns similar to those observed for the XGBoost model. Corresponding correlation plots (**Fig 5e**) validate these findings, demonstrating similar directional relationships and highlighting the strong influence of these features on TUG prediction.

## Discussion

This study introduces a novel two-stage machine learning pipeline to predict Berg Balance Scale (BBS) and Timed Up and Go (TUG) scores, employing binary classification for feature selection to manage high-dimensionality (528 features, n=33) and reduce overfitting, while identifying biomarkers distinguishing stroke participants from healthy controls [35]. By assessing seven models across three seeds (42, 100, 123), the classification task ensured robustness, with XGBoost and Elastic Net achieving perfect Test ROC AUC, justifying their selection for regression to maintain consistent feature importance [98]. The classification step yielded 15 key features, including BBS and TUG, showing that these outcomes differentiate stroke from healthy controls. Successive machine learning-based regression analysis was performed with the remaining 13 features to predict BBS and TUG. We found that a combination of CMC and force platform-based measures predicts both BBS and TUG in stroke and healthy participants, suggesting that these features capture mechanisms shared across balance and functional mobility. These features included Tibialis Anterior delta-band CMC on the affected side of stroke or the dominant side of healthy control, and the medio-lateral COP displacement. We also found that the asymmetry in soleus theta-band CMC between the two lower limbs predicts BBS. Finally, we found that the asymmetry in biceps femoris beta band CMC between the two lower limbs, and the rectus femoris beta band CMC were unique features that predicted TUG test scores. These findings provide objective, clinically meaningful biomarkers that may be sensitive to subtle changes in balance behavior following an intervention.

### Neuromechanical Features that Capture both Balance and Functional Mobility

Our machine learning based regression models found tibialis anterior delta-band coherence as one of the top three predictors of BBS and TUG. Mainly, higher TA delta-band coherence was associated with higher BBS scores and faster TUG completion times, indicating better balance and functional mobility. The TA muscle, an ankle dorsiflexor, is crucial for stability, especially in the anterior-posterior direction. It helps maintain equilibrium against gravitational perturbations during static and dynamic balance (BBS) and provides foot clearance and forward propulsion in transitional movements and walking (TUG). Impaired TA function can compromise these abilities, as seen with foot drop common post-stroke [79,106,107]. The Cz-TA coherence measure provides oscillatory synchrony between the sensorimotor cortical region and the TA muscle involved in balance and functional mobility. The oscillations in the delta frequency band are important for attentional modulation, error detection, and adaptive postural responses [13,108,109]. The lower frequency rhythms integrate multisensory inputs, including proprioceptive and vestibular, via feedback loops involving the corticospinal tract and basal ganglia-thalamic circuits, into coherent information to assist in counteracting sway and ensure stability when balance is perturbed [110–113]. Additionally, TA delta-band oscillatory synchrony may also reflect anticipatory postural adjustments important for both BBS and TUG, which require shared motor control processes. [114,115]. When task difficulty was higher (e.g., medium difficulty continuous balance task condition), TA delta-band coherence significantly predicted both BBS and TUG, suggesting its ability to capture balance and functional mobility under more challenging/dynamic balance conditions. Conversely, lower TA delta-band coherence in stroke participants might suggest desynchronization of the cortical muscle signaling, affecting the use of sensory information for precise modulation of the TA muscle. The resulting increase in sway variability and compensatory overactivation of unaffected limbs reduces BBS and prolongs TUG [13,116]. Together, these findings suggest that the TA delta band coherence may be sensitive to subtle recovery changes [2,18] and is a promising objective marker of balance and functional mobility following stroke.

The RMS center of pressure measure, which measures sway amplitude in the medio-lateral (ML) direction over time, was another common predictor of Balance Scale (BBS) and Timed Up and Go (TUG) outcomes in our study. Higher RMS COP_ML_ during the most challenging balance task condition was associated with poor scores on the BBS and TUG tests. Although the continuous balance task was to be performed in the sagittal plane, the participants were allowed unrestricted movements in the medio-lateral direction but without changing the base of support. A response to large balance disturbances in the sagittal plane requires the engagement of powerful muscles at the hips to stabilize the body’s center of mass and control side-to-side movement. Because of weak or poorly coordinated hip abductor muscles following stroke [117,118], there is a greater reliance on the ankle strategy, which is insufficient to stabilize the body’s lateral center of mass during large balance disturbances, increasing medio-lateral sway variability [32,119]. The tendency of our participants to show increased lateral sway under dynamic balance situations might have increased energy cost, reduced movement efficiency, and slowed functional mobility [29,30,33,120]. Our findings suggest that RMS COP_ML_ may be a valuable marker for tracking balance and functional mobility impairments in stroke rehabilitation [26,121,122].

### Unique Predictors of Balance

CMC asymmetry of the soleus muscle in the theta band during the medium condition served as a predictor of Berg Balance Scale (BBS) scores but not TUG. Soleus asymmetry index measures the differences in synchronized brain-muscle signaling (CMC) between the affected and less affected in stroke and dominant-non-dominant soleus muscles. Higher asymmetry in the soleus theta-band CMC between the two sides was associated with lower scores on the Berg Balance Scale, reflecting imbalance in the neural control during balance tasks. The soleus muscle primarily performs ankle plantarflexion, helping resist forward sway and maintain upright posture using fixed-support strategies [123,124]. The soleus muscle is composed of slow-twitch, fatigue-resistant (type I) fibers [125], specialized for sustained, low-intensity contractions and antigravity control, suitable for balance tasks such as those performed during the Berg Balance test. Asymmetrical recruitment of these fibers leads to over-reliance on the less-affected side in stroke participants and the non-dominant side in healthy controls, increasing sway and instability [126,127]. The observation of higher soleus CMC asymmetry in the theta-band frequency (∼4-8 Hz) in stroke participants suggests impaired attention and sensorimotor integration, supporting postural adjustments necessary for maintaining a steady stance [126,128–131]. Overall, soleus theta-band CMC asymmetry complements other markers in multimodal models, assisting in tracking BBS-specific impairments [28].

### Unique Predictors of Functional Mobility

Asymmetry in the beta-band CMC for the biceps femoris muscle during challenging perturbation tasks emerged as a specific predictor of Timed Up and Go (TUG) performance, showing a positive correlation where greater asymmetry was linked to prolonged TUG times. Biceps femoris, a key muscle for knee flexion and hip extension, stabilizes the body as well as provides propulsive force during dynamic tasks such as walking and turning phases [132]. The asymmetry in the coupling between the sensorimotor region and the biceps femoris muscle reflects an imbalance in the neural control during dynamic balance tasks, which can hinder functional mobility [133,134]. The beta-band rhythm (∼12–30 Hz) contributes to precise motor adjustments and coordination, supporting stability and efficient movement during complex functional mobility tasks [135,136]. Heightened asymmetry under challenging balance conditions indicates ineffective compensatory strategies that affect dynamic balance control, crucial for functional mobility [135,137]. Thus, biceps femoris beta-band CMC asymmetry serves as a sensitive marker for tracking functional mobility improvements in stroke [28].

Rectus femoris beta-band coherence during challenging perturbation tasks and the modulation of RF CMC across task difficulty on the less affected side in stroke and the non-dominant side in healthy participants emerged as significant predictors of Timed Up and Go (TUG) performance, both demonstrating negative correlations—indicating that higher RF beta-band coherence and its greater modulation were associated with reduced TUG times, thus better functional mobility. The rectus femoris muscle plays a critical role in knee extension and hip flexion, essential for forward propulsion and stability during dynamic functional mobility [138]. In addition, both these markers reflect synchronized cortico-muscular communication in the beta frequency range (∼12–30 Hz), which facilitates motor adjustments and sensorimotor integration during movement [135,136,139]. During challenging balance tasks, higher beta coherence may indicate more effective corticospinal engagement to maintain stability under perturbations [140,141]. Similarly, greater modulation of coherence across varying task difficulty may suggest better neural and behavioral adaptability to changing demands, reflecting preserved or compensatory motor control mechanisms [142,143]. This adaptability likely enhances dynamic balance control, thus reducing TUG times. Importantly, because the rectus femoris is a biarticular muscle involved in both propulsion and postural stabilization, greater beta-band coherence and modulation may also reflect energy-efficient motor strategies that minimize metabolic cost during transitions such as sit-to-stand and turning,

improving overall functional mobility [135,144]. Collectively, these findings highlight rectus femoris beta-band coherence and its task-related modulation as promising neurophysiological biomarkers for functional mobility, complementing other measures in multimodal models for tracking functional recovery in stroke rehabilitation [28,145]. However, while reliance on the less affected side may support functional mobility, it can also reflect compensatory strategies that may neglect recovery of the more affected limb. Therefore, these markers may be viewed as indicators of functional adaptation rather than complete restoration of bilateral motor control.

### Integrated Insights for Rehabilitation

Our study identified CMC and force platform-based predictors that reveal distinct mechanisms for balance and functional mobility based on muscle type, frequency bands, and task complexity. Distal muscles like the tibialis anterior and soleus, by utilizing low-frequency oscillatory synchrony, support fixed-support ankle strategies crucial during the Berg Balance test. On the other hand, proximal muscles like the rectus femoris and biceps femoris, through high frequency coherence, enable change-in-support (hip) strategies essential during the Timed-up and Go test [146–148]. This shift from low-frequency (delta–theta) ankle control to higher-frequency (beta) hip control with increasing task demands reflects compensatory mechanisms after stroke. Combining these CMC measures with center-of-pressure metrics in multimodal approaches provides objective biomarkers beyond subjective clinical assessments, providing insights into how neural and biomechanical systems coordinate to maintain stability and mobility. This can help tailor rehabilitation to improve balance and functional mobility, boost independence, and monitor progress effectively [8]. Larger studies are needed to validate and expand these applications to support stroke rehabilitation.

### Limitations

This study’s small sample size (n=33, 18 stroke patients, 15 healthy controls) limits the generalizability of findings, particularly for regression models, where negative adjusted R² values on the test set (e.g., BBS: −0.8816 for Elastic Net; TUG: −6.8216 for XGBoost) may suggest overfitting due to limited test samples (n=9) despite robust classification performance [98]. The high-dimensional dataset (528 features) required a classification-based feature selection pipeline, which, while effective in reducing dimensionality to 13 predictors, may have excluded potentially relevant features [35]. In addition, the reliance on a single seed (42) for regression, compared to three seeds for classification, may reduce the robustness of predictive performance estimates. The use of KNN imputation for missing values (23 columns with significant deviations, p < 0.05) may introduce potential bias, particularly if group-specific patterns were not fully preserved [95]. Finally, the lack of validation on a separate dataset limits the application of identified predictors to a broader stroke/elderly population, thus necessitating further studies to confirm their predictive/marker value.

### Conclusion

This study identifies clinically relevant objective markers for predicting BBS and TUG performance in stroke rehabilitation by integrating CMC and COP measures via machine learning approaches. Predictors such as tibialis anterior delta-band coherence and medio-lateral COP sway reveal the control strategies shared across BBS and TUG. Muscle- and frequency-specific predictors, such as soleus theta CMC asymmetry for BBS and biceps femoris/rectus femoris beta-band CMC for TUG, highlight distinct neuromuscular control strategies. These objective markers offer potentially greater sensitivity than traditional assessments, enabling personalized interventions to improve outcomes and reduce falls. Future larger-scale studies will enhance their clinical applications for tracking functional recovery.

## Data Availability

The data that support the findings of this study are available from the corresponding author upon request, conditional upon participant consent regarding data sharing. Raw EEG, EMG, and force platform data contain sensitive participant information and are not publicly available to protect privacy, but de-identified processed data and codes may be provided for verification purposes.

## Acknowledgements

This study was partially supported by a grant from the National Institutes of Health Eunice Kennedy Shriver National Institute of Child Health and Human Development R25HD106896 and the National Science Foundation IUCRC BRAIN Center awards 2137255 and 2137272 to PJP.

## REFERENCES

[1] Yang C, Ghaedi B, Campbell TM, Rutkowski N, Finestone H. Predicting Falls Using the Stroke Assessment of Fall Risk Tool. PM and R 2021;13:274–81. 10.1002/pmrj.12434.

[2] Li J, Zhong D, Ye J, He M, Liu X, Zheng H, et al. Rehabilitation for balance impairment in patients after stroke: a protocol of a systematic review and network meta-analysis. BMJ Open 2019;9:26844. 10.1136/BMJOPEN-2018-026844.

[3] Denissen S, Staring W, Kundel D, Pickering RM, Lennon S, Geurts ACH, et al. Cochrane Library Cochrane Database of Systematic Reviews Interventions for preventing falls in people aer stroke (Review) 2019. 10.1002/14651858.CD008728.pub3.

[4] Alenazi AM, Alshehri MM, Alothman S, Rucker J, Dunning K, D’Silva LJ, et al. Functional Reach, Depression Scores, and Number of Medications Are Associated With Number of Falls in People With Chronic Stroke. PM and R 2018;10:806–16. 10.1016/j.pmrj.2017.12.005.

[5] Tasseel-Ponche S, Yelnik AP, Bonan I V. Motor strategies of postural control after hemispheric stroke. Neurophysiologie Clinique 2015;45:327–33. 10.1016/j.neucli.2015.09.003.

[6] Nagarkar A, Kulkarni S. Association between daily activities and fall in older adults: an analysis of longitudinal ageing study in India (2017–18). BMC Geriatr 2022;22. 10.1186/s12877-022-02879-x.

[7] Pua YH, Ong PH, Clark RA, Matcher DB, Lim ECW. Falls efficacy, postural balance, and risk for falls in older adults with falls-related emergency department visits: prospective cohort study. BMC Geriatr 2017;17:291. 10.1186/S12877-017-0682-2.

[8] Wade E, Winstein CJ. Virtual reality and robotics for stroke rehabilitation: where do we go from here? Top Stroke Rehabil 2011;18:685–700. 10.1310/TSR1806-685.

[9] Mao H-F, Hsueh I-P, Tang P-F, Sheu C-F, Hsieh C-L. Analysis and Comparison of the Psychometric Properties of Three Balance Measures for Stroke Patients. 2002.

[10] Podsiadlo D, Richardson S. The Timed “Up & Go”: A Test of Basic Functional Mobility for Frail Elderly Persons. vol. 39. 1991.

11. Blum L, Korner-Bitensky N. Usefulness_of_the_Berg_Balance 2008.

[12] Tyson SF, Connell LA. How to measure balance in clinical practice. A systematic review of the psychometrics and clinical utility of measures of balance activity for neurological conditions. Clin Rehabil 2009;23:824–40. 10.1177/0269215509335018.

[13] Kukkar KK, Rao N, Huynh D, Shah S, Contreras-Vidal JL, Parikh PJ. Context-dependent reduction in corticomuscular coupling for balance control in chronic stroke survivors. Exp Brain Res 2024.

[14] Bruyneel AV, Mesure S, Reinmann A, Sordet C, Venturelli P, Feldmann I, et al. Validity and reliability of center of pressure measures to quantify trunk control ability in individuals after stroke in subacute phase during unstable sitting test. Heliyon 2022;8:e10891. 10.1016/J.HELIYON.2022.E10891.

[15] Conway BA, Halliday DM, Farmer SF, Shahani U, Maas P, Weir AI, et al. Synchronization between motor cortex and spinal motoneuronal pool during the performance of a maintained motor task in man. J Physiol 1995;489:917. 10.1113/JPHYSIOL.1995.SP021104.

[16] Bonanno M, Maggio MG, De Pasquale P, Ciatto L, Lombardo Facciale A, De Francesco M, et al. The Potential Effects of Sensor-Based Virtual Reality Telerehabilitation on Lower Limb Function in Patients with Chronic Stroke Facing the COVID-19 Pandemic: A Retrospective Case-Control Study. Med Sci (Basel) 2025;13. 10.3390/MEDSCI13020065.

[17] Petersen TH, Willerslev-Olsen M, Conway BA, Nielsen JB. The motor cortex drives the muscles during walking in human subjects. Journal of Physiology 2012;590:2443–52. 10.1113/jphysiol.2012.227397.

[18] Xu R, Zhang H, Shi X, Liang J, Wan C, Ming D. Lower-Limb Motor Assessment With Corticomuscular Coherence of Multiple Muscles During Ankle Dorsiflexion After Stroke. IEEE Transactions on Neural Systems and Rehabilitation Engineering 2023;31:160–8. 10.1109/TNSRE.2022.3217571.

[19] Chen IH, Yang YR, Lu CF, Wang RY. Novel gait training alters functional brain connectivity during walking in chronic stroke patients: A randomized controlled pilot trial. J Neuroeng Rehabil 2019;16:1–14. 10.1186/S12984-019-0503-2/TABLES/4.

[20] Hasui N, Mizuta N, Matsunaga A, Higa Y, Sato M, Nakatani T, et al. Association of gait recovery with intramuscular coherence of the Vastus medialis muscle during assisted gait in subacute stroke. Neurological Sciences 2025;46:3735–45. 10.1007/S10072-025-08138-2/FIGURES/3.

[21] Bao SC, Sun R, Tong RKY. Pedaling Asymmetry Reflected by Bilateral EMG Complexity in Chronic Stroke. Entropy 2024, Vol 26, Page 538 2024;26:538. 10.3390/E26070538.

[22] Park S-K, Yang D-J, Kim J-H, Jeong Y-S. Correlation Between BBS, TUG and Lower Extremity Muscle Activity during Semi-Squat in Stroke Patients. Journal of the Korean Academy of Clinical Electrophysiology 2011;9:7–12. 10.5627/KACE.2011.9.2.007.

[23] Kwong PWH, Ng GYF, Chung RCK, Ng SSM. Bilateral Transcutaneous Electrical Nerve Stimulation Improves Lower-Limb Motor Function in Subjects With Chronic Stroke: A Randomized Controlled Trial. J Am Heart Assoc 2018;7. 10.1161/JAHA.117.007341.

[24] Jeon HJ, Hwang BY. Effect of bilateral lower limb strengthening exercise on balance and walking in hemiparetic patients after stroke: a randomized controlled trial. J Phys Ther Sci 2018;30:277–81. 10.1589/JPTS.30.277.

[25] Quijoux F, Vienne-Jumeau A, Bertin-Hugault F, Lefèvre M, Zawieja P, Vidal PP, et al. Center of pressure characteristics from quiet standing measures to predict the risk of falling in older adults: A protocol for a systematic review and meta-analysis. Syst Rev 2019;8. 10.1186/S13643-019-1147-9,.

[26] Quijoux F, Vienne-Jumeau A, Bertin-Hugault F, Zawieja P, Lefèvre M, Vidal PP, et al. Center of pressure displacement characteristics differentiate fall risk in older people: A systematic review with meta-analysis. Ageing Res Rev 2020;62. 10.1016/j.arr.2020.101117.

[27] Cheng PT, Liaw MY, Wong MK, Tang FT, Lee MY, Lin PS. The sit-to-stand movement in stroke patients and its correlation with falling. Arch Phys Med Rehabil 1998;79:1043–6. 10.1016/S0003-9993(98)90168-X.

[28] Alhasan HS, Wheeler PC, Fong DTP. Application of Interactive Video Games as Rehabilitation Tools to Improve Postural Control and Risk of Falls in Prefrail Older Adults. Cyborg and Bionic Systems 2021;2021. 10.34133/2021/9841342.

[29] Karunakaran KK, Pamula S, Tendolkar P, Chen P, Suviseshamuthu ES. Preliminary Validation of an Objective Fall-Risk Assessment for Individuals with Stroke. Proceedings of the Annual International Conference of the IEEE Engineering in Medicine and Biology Society, EMBS, Institute of Electrical and Electronics Engineers Inc.; 2024. 10.1109/EMBC53108.2024.10782236.

[30] Onursal Kilinç Ö, Ayvat E, Ayvat F, Kilinç M. Association of posturography with clinical measures in balance rehabilitation of ataxic patients. International Journal of Rehabilitation Research 2021;44:256–61. 10.1097/MRR.0000000000000481.

[31] Lichtenstein MJ, Burger MC, Shields SL, Shiavi RG. Comparison of Biomechanics Platform Measures of Balance and Videotaped Measures of Gait With a Clinical Mobility Scale in Elderly Women. J Gerontol 1990;45:M49–54. 10.1093/GERONJ/45.2.M49.

[32] Allum JHJ, Adkin AL. Allum & Adkin, 2003. Audiol Neurootol 2003.

[33] Moon Y, Kim M-S, Choi J-D. Correlation between Weight Bearing Ratio and Functional Level for Development of Pressure Sensor Biofeedback in Stroke Patient. Journal of the Korean Society of Physical Medicine 2014;9:315–24. 10.13066/kspm.2014.9.3.315.

[34] Sawacha Z, Carraro E, Contessa P, Guiotto A, Masiero S, Cobelli C. Relationship between clinical and instrumental balance assessments in chronic post-stroke hemiparesis subjects. J Neuroeng Rehabil 2013;10. 10.1186/1743-0003-10-95.

[35] Saeys Y, Inza I, Larrañaga P. A review of feature selection techniques in bioinformatics. Bioinformatics 2007;23:2507–17. 10.1093/BIOINFORMATICS/BTM344.

[36] Theng D, Bhoyar KK. Feature selection techniques for machine learning: a survey of more than two decades of research. Knowl Inf Syst 2024;66:1575–637. 10.1007/S10115-023-02010-5/TABLES/6.

[37] Bränström R, Bränström R, Pernow Y, Bränström R, Nilsson IL. Prediction of cognitive response to surgery in elderly patients with primary hyperparathyroidism. BJS Open 2021;5. 10.1093/bjsopen/zraa029.

[38] Trzepacz PT, Hochstetler H, Wang S, Walker B, Saykin AJ. Relationship between the Montreal Cognitive Assessment and Mini-mental State Examination for assessment of mild cognitive impairment in older adults. BMC Geriatr 2015;15. 10.1186/s12877-015-0103-3.

[39] Cohen HS, Kimball KT. Usefulness of some current balance tests for identifying individuals with disequilibrium due to vestibular impairments. J Vestib Res 2008;18:295–303.

[40] Wood SJ, Paloski WH, Clark JB. Assessing Sensorimotor Function Following ISS with Computerized Dynamic Posturography. Aerosp Med Hum Perform 2015;86:A45–53. 10.3357/AMHP.EC07.2015.

[41] Berg KO, Wood-Dauphinee SL, Williams JI, Maki B. Measuring balance in the elderly: Validation of an instrument. Canadian Journal of Public Health, vol. 83, 1992.

[42] Podsiadlo JD, Bscpt S, Richardson MDJ. The Timed ‘Up & Go’: A Test of Basic Functional Mobilitv for Frail Elderlv Persons. vol. 39. 1991.

[43] Rasman BG, Forbes PA, Tisserand R, Blouin JS. Sensorimotor manipulations of the balance control loop-beyond imposed external perturbations. Front Neurol 2018;9. 10.3389/fneur.2018.00899.

[44] Goel R, Nakagome S, Rao N, Paloski WH, Contreras-Vidal JL, Parikh PJ. Fronto-Parietal Brain Areas Contribute to the Online Control of Posture during a Continuous Balance Task. Neuroscience 2019;413:135–53. 10.1016/j.neuroscience.2019.05.063.

[45] Ozdemir RA, Contreras-Vidal JL, Paloski WH. Cortical control of upright stance in elderly. Mech Ageing Dev 2018;169:19–31. 10.1016/j.mad.2017.12.004.

[46] Ozdemir RA, Contreras-Vidal JL, Lee BC, Paloski WH. Cortical activity modulations underlying age-related performance differences during posture–cognition dual tasking. Exp Brain Res 2016;234:3321–34. 10.1007/s00221-016-4730-5.

[47] Luu TP, Brantley JA, Nakagome S, Zhu F, Contreras-Vidal JL. Electrocortical correlates of human level-ground, slope, and stair walking. PLoS One 2017;12. 10.1371/journal.pone.0188500.

[48] Luu TP, He Y, Brown S, Nakagame S, Contreras-Vidal JL. Gait adaptation to visual kinematic perturbations using a real-time closed-loop brain–computer interface to a virtual reality avatar. J Neural Eng 2016;13:036006. 10.1088/1741-2560/13/3/036006.

[49] Mohapatra S, Kukkar KK, Aruin AS. Support surface related changes in feedforward and feedback control of standing posture. Journal of Electromyography and Kinesiology 2014;24. 10.1016/j.jelekin.2013.10.015.

[50] Santos MJ, Kanekar N, Aruin AS. The role of anticipatory postural adjustments in compensatory control of posture: 1. Electromyographic analysis. Journal of Electromyography and Kinesiology 2010;20:388–97. 10.1016/j.jelekin.2009.06.006.

[51] Neurocom. Clinical Interpretation Guide - Computerized Dynamic Posturography. 2009.

[52] Goel R, Nagakome S, Parikh P. Goel 2021 2021.

[53] Breniere Y. Why we walk the way we do. J Mot Behav 1996;28:291–8. 10.1080/00222895.1996.10544598.

[54] Caron O, Faure B, Breniere Y. Estimating the centre of gravity of the body on the basis of the centre of pressure in standing posture. J Biomech 1997;30:1169–71. 10.1016/s0021-9290(97)00094-8.

[55] Lafond D, Duarte M, Prince F. Comparison of three methods to estimate the center of mass during balance assessment. J Biomech 2004;37:1421–6. 10.1016/s0021-9290(03)00251-3.

[56] Prieto TE, Myklebust JB, Hoffmann RG, Lovett EG, Myklebust BM. Measures of postural steadiness: Differences between healthy young and elderly adults. IEEE Trans Biomed Eng 1996;43:956–66. 10.1109/10.532130.

[57] Geurts ACH, Nienhuis B, Maartenskliniek S, Mulder T. Article in Archives of Physical Medicine and Rehabilitation. 1993.

[58] Donath L, Roth R, Zahner L, Faude O. Testing single and double limb standing balance performance: comparison of COP path length evaluation between two devices. Gait Posture 2012;36:439–43. 10.1016/j.gaitpost.2012.04.001.

[59] Forth KE, Fiedler MJ, Paloski WH. Estimating functional stability boundaries for bipedal stance. Gait Posture 2011;33:715–7. 10.1016/j.gaitpost.2010.12.024.

[60] Ozdemir RA, Contreras-Vidal JL, Paloski WH. Cortical control of upright stance in elderly. Mech Ageing Dev 2018;169:19–31. 10.1016/j.mad.2017.12.004.

[61] Ozdemir RA, Pourmoghaddam A, Paloski WH. Sensorimotor posture control in the blind: superior ankle proprioceptive acuity does not compensate for vision loss. Gait Posture 2013;38:603–8. 10.1016/j.gaitpost.2013.02.003.

[62] Schubert P, Kirchner M. Ellipse area calculations and their applicability in posturography. Gait Posture 2014;39:518–22. 10.1016/J.GAITPOST.2013.09.001.

63. Duarte M, Freitas SMSF. Revision of posturography based on force plate for balance evaluation Revisão sobre posturografia baseada em plataforma de força para avaliação do equilíbrio n.d.

[64] Magruder RD, Kukkar KK, Contreras-Vidal JL, Parikh PJ. Cross-Task Differences in Frontocentral Cortical Activations for Dynamic Balance in Neurotypical Adults. Sensors (Basel) 2024;24. 10.3390/S24206645.

[65] Dadfar M, Kukkar KK, Parikh PJ. Reduced parietal to frontal functional connectivity for dynamic balance in late middle-to-older adults. Exp Brain Res 2025;243. 10.1007/S00221-025-07070-3.

[66] Kilicarslan A, Grossman RG, Contreras-Vidal JL. A robust adaptive denoising framework for real-time artifact removal in scalp EEG measurements. J Neural Eng 2016;13:026013. 10.1088/1741-2560/13/2/026013.

[67] De Cheveigné A, Nelken I. Primer Filters: When, Why, and How (Not) to Use Them. Neuron 2019;102:280–93. 10.1016/j.neuron.2019.02.039.

[68] Bigdely-Shamlo N, Mullen T, Kothe C, Su K-M, Robbins KA. The PREP pipeline: standardized preprocessing for large-scale EEG analysis. Front Neuroinform 2015;9:16. 10.3389/fninf.2015.00016.

[69] Delorme A, Makeig S. EEGLAB: An open source toolbox for analysis of single-trial EEG dynamics including independent component analysis. J Neurosci Methods 2004;134:9–21. 10.1016/j.jneumeth.2003.10.009.

[70] Mullen T, Kothe C, Chi YM, Ojeda A, Kerth T, Makeig S, et al. Real-time modeling and 3D visualization of source dynamics and connectivity using wearable EEG. Proceedings of the Annual International Conference of the IEEE Engineering in Medicine and Biology Society, EMBS, 2013, p. 2184–7. 10.1109/EMBC.2013.6609968.

[71] Artoni F, Fanciullacci C, Bertolucci F, Panarese A, Makeig S, Micera S, et al. Unidirectional brain to muscle connectivity reveals motor cortex control of leg muscles during stereotyped walking. Neuroimage 2017;159:403–16. 10.1016/j.neuroimage.2017.07.013.

[72] Chang CY, Hsu SH, Pion-Tonachini L, Jung TP. Evaluation of Artifact Subspace Reconstruction for Automatic Artifact Components Removal in Multi-Channel EEG Recordings. IEEE Trans Biomed Eng 2020;67:1114–21. 10.1109/TBME.2019.2930186.

[73] Palmer JA, Kreutz-Delgado K, Makeig S. AMICA: An adaptive mixture of independent component analyzers with shared components 2011.

[74] Delorme A, Palmer J, Onton J, Oostenveld R, Makeig S. Independent EEG sources are dipolar. PLoS One 2012;7:e30135.

[75] Leutheuser H, Gabsteiger F, Hebenstreit F, Reis P, Lochmann M, Eskofier B. Comparison of the AMICA and the InfoMax algorithm for the reduction of electromyogenic artifacts in EEG data. 2013 35th Annual International Conference of the IEEE Engineering in Medicine and Biology Society (EMBC), IEEE; 2013, p. 6804–7.

[76] Delorme A, Mullen T, Kothe C, Acar ZA, Bigdely-Shamlo N, Vankov A, et al. EEGLAB, SIFT, NFT, BCILAB, and ERICA: new tools for advanced EEG processing. Comput Intell Neurosci 2011;2011:10.

[77] Sipp AR, Gwin JT, Makeig S, Ferris DP. Loss of balance during balance beam walking elicits a multifocal theta band electrocortical response. J Neurophysiol 2013;110:2050–60.

78. Neuper C, Pfurtscheller G. Evidence for distinct beta resonance frequencies in human EEG related to specific sensorimotor cortical areas. 2001.

[79] Ushiyama J, Masakado Y, Fujiwara T, Tsuji T, Hase K, Kimura A, et al. Contraction level-related modulation of corticomuscular coherence differs between the tibialis anterior and soleus muscles in humans. J Appl Physiol 2012;112:1258–67. 10.1152/japplphysiol.01291.2011.-The.

[80] Grinsted A, Moore JC, Jevrejeva S. Nonlinear Processes in Geophysics Application of the cross wavelet transform and wavelet coherence to geophysical time series. vol. 11. 2004.

[81] Witham CL, Riddle CN, Baker MR, Baker SN. Contributions of descending and ascending pathways to corticomuscular coherence in humans. Journal of Physiology 2011;589:3789– 800. 10.1113/jphysiol.2011.211045.

[82] Baker MR, Baker SN. The effect of diazepam on motor cortical oscillations and corticomuscular coherence studied in man. Journal of Physiology 2003;546:931–42. 10.1113/jphysiol.2002.029553.

[83] Mima T, Hallett M. Electroencephalographic analysis of cortico-muscular coherence: reference effect, volume conduction and generator mechanism. 1999.

[84] Chen YT, Li S, Magat E, Zhou P, Li S. Motor Overflow and Spasticity in Chronic Stroke Share a Common Pathophysiological Process: Analysis of Within-Limb and Between-Limb EMG-EMG Coherence. Front Neurol 2018;9. 10.3389/FNEUR.2018.00795.

[85] Grosse P, Brown P. Acoustic startle evokes bilaterally synchronous oscillatory EMG activity in the healthy human. J Neurophysiol 2003;90:1654–61. 10.1152/JN.00125.2003.

[86] Van Asseldonk EHF, Campfens SF, Verwer SJF, Van Putten MJAM, Stegeman DF. Reliability and agreement of intramuscular coherence in tibialis anterior muscle. PLoS One 2014;9. 10.1371/JOURNAL.PONE.0088428.

[87] Gao Z, Lv S, Ran X, Wang Y, Xia M, Wang J, et al. Influencing factors of corticomuscular coherence in stroke patients. Front Hum Neurosci 2024;18. 10.3389/fnhum.2024.1354332.

[88] Mima T, Toma K, Koshy B, Hallett M. Coherence Between Cortical and Muscular Activities After Subcortical Stroke. 2001.

[89] Krauth R, Schwertner J, Vogt S, Lindquist S, Sailer M, Sickert A, et al. Cortico-Muscular Coherence Is Reduced Acutely Post-stroke and Increases Bilaterally During Motor Recovery: A Pilot Study. Front Neurol 2019;10. 10.3389/FNEUR.2019.00126.

[90] Hadamus A, Gulatowska M, Ferenc A, Shahnazaryan K, Brzuszkiewicz-Kuźmicka G, Błażkiewicz M. Influence of leg dominance on the symmetry in body balance measurements. Physical Activity Review 2025;13:88–96. 10.16926/par.2025.13.08.

[91] Kristeva R, Patino L, Omlor W. Beta-range cortical motor spectral power and corticomuscular coherence as a mechanism for effective corticospinal interaction during steady-state motor output. Neuroimage 2007;36:785–92. 10.1016/J.NEUROIMAGE.2007.03.025.

[92] Hari R, Salenius S. Rhythmical corticomotor communication. Neuroreport 1999;10.

[93] Gudmundsson S, Runarsson TP, Sigurdsson S, Eiriksdottir G, Johnsen K. Reliability of quantitative EEG features. Clinical Neurophysiology 2007;118:2162–71. 10.1016/j.clinph.2007.06.018.

[94] von Carlowitz-Ghori K, Bayraktaroglu Z, Hohlefeld FU, Losch F, Curio G, Nikulin V V. Corticomuscular coherence in acute and chronic stroke. Clinical Neurophysiology 2014;125:1182–91. 10.1016/j.clinph.2013.11.006.

[95] Troyanskaya O, Cantor M, Sherlock G, Brown P, Hastie T, Tibshirani R, et al. Missing value estimation methods for DNA microarrays. Bioinformatics 2001;17:520–5. 10.1093/BIOINFORMATICS/17.6.520,.

[96] Little RJA., Rubin DB. Statistical analysis with missing data 2020:449.

[97] Breiman L. Random forests. Mach Learn 2001;45:5–32. 10.1023/A:1010933404324/METRICS.

[98] Hastie T, Tibshirani R, Friedman J. The Elements of Statistical Learning 2009. 10.1007/978-0-387-84858-7.

99. Kohavi R. A Study of Cross-Validation and Bootstrap for Accuracy Estimation and Model Selection. 1995.

[100] Pedregosa FABIANPEDREGOSA F, Michel V, Grisel OLIVIERGRISEL O, Blondel M, Prettenhofer P, Weiss R, et al. Scikit-learn: Machine Learning in Python. Journal of Machine Learning Research 2011;12:2825–30.

[101] Ca BU, Fr YG. No Unbiased Estimator of the Variance of K-Fold Cross-Validation. Journal of Machine Learning Research 2004;5:1089–105.

[102] James G, Witten D, Hastie T, Tibshirani R. An Introduction to Statistical Learning 2021. 10.1007/978-1-0716-1418-1.

[103] Fawcett T. An introduction to ROC analysis. Pattern Recognit Lett 2006;27:861–74. 10.1016/J.PATREC.2005.10.010.

104. Lundberg SM, Allen PG, Lee S-I. A Unified Approach to Interpreting Model Predictions. 2017.

[105] McInnes L, Healy J, Melville J. UMAP: Uniform Manifold Approximation and Projection for Dimension Reduction 2018.

[106] He R, Dong Y, Li Y, Zheng M, Peng S, Tong RKY, et al. Therapeutic and orthotic effects of an adaptive functional electrical stimulation system on gait biomechanics in participants with stroke. J Neuroeng Rehabil 2025;22:1–12. 10.1186/S12984-025-01577-0/TABLES/2.

[107] Ramsay JW, Wessel MA, Buchanan TS, Higginson JS. Poststroke Muscle Architectural Parameters of the Tibialis Anterior and the Potential Implications for Rehabilitation of Foot Drop. Stroke Res Treat 2014;2014:948475. 10.1155/2014/948475.

[108] Novembre G, Pawar VM, Kilintari M, Bufacchi RJ, Guo Y, Rothwell JC, et al. The effect of salient stimuli on neural oscillations, isometric force, and their coupling. Neuroimage 2019;198:221–30. 10.1016/j.neuroimage.2019.05.032.

[109] Novembre G, Pawar VM, Bufacchi RJ, Kilintari M, Srinivasan M, Rothwell JC, et al. Saliency detection as a reactive process: Unexpected sensory events evoke corticomuscular coupling. Journal of Neuroscience 2018;38:2385–97. 10.1523/JNEUROSCI.2474-17.2017.

[110] Legrand T, Mongold SJ, Muller L, Naeije G, Ghinst M Vander, Bourguignon M. Cortical tracking of postural sways during standing balance. Sci Rep 2024;14. 10.1038/S41598-024-81865-2.

[111] Gelormini C, Guerrini L, Pescaglia F, Aubonnet R, Jónsson H, Petersen H, et al. Assessing Brain Network Dynamics During Postural Control Task Using EEG Microstates. Brain Topogr 2025;38. 10.1007/S10548-025-01119-W.

[112] Ertl M, Zu Eulenburg P, Woller M, Dieterich M. The role of delta and theta oscillations during ego-motion in healthy adult volunteers. Exp Brain Res 2021;239:1073–83. 10.1007/S00221-020-06030-3.

[113] Hua A, Wang G, Bai J, Hao Z, Liu J, Meng J, et al. Nonlinear dynamics of postural control system under visual-vestibular habituation balance practice: evidence from EEG, EMG and center of pressure signals. Front Hum Neurosci 2024;18:1371648. 10.3389/FNHUM.2024.1371648/BIBTEX.

[114] Berg WP. Advances in the Study of Anticipatory Postural Adjustments. Brain Sciences 2024, Vol 14, Page 1219 2024;14:1219. 10.3390/BRAINSCI14121219.

[115] Farinelli V, Bolzoni F, Marchese SM, Esposti R, Cavallari P. A Novel Viewpoint on the Anticipatory Postural Adjustments During Gait Initiation. Front Hum Neurosci 2021;15:709780. 10.3389/FNHUM.2021.709780/BIBTEX.

[116] Paul T, Cieslak M, Hensel L, Wiemer VM, Grefkes C, Grafton ST, et al. The role of corticospinal and extrapyramidal pathways in motor impairment after stroke. Brain Commun 2022;5. 10.1093/BRAINCOMMS/FCAC301.

[117] Darak V, Karthikbabu S. Lower limb motor function and hip muscle weakness in stroke survivors and their relationship with pelvic tilt, weight-bearing asymmetry, and gait speed: A cross-sectional study. Curr J Neurol 2020;19:1–7. 10.18502/IJNL.V19I1.3275.

[118] Sánchez N, Acosta AM, López-Rosado R, Dewald JPA. Neural Constraints Affect the Ability to Generate Hip Abduction Torques When Combined With Hip Extension or Ankle Plantarflexion in Chronic Hemiparetic Stroke. Front Neurol 2018;9. 10.3389/FNEUR.2018.00564.

[119] De Haart M, Geurts AC, Huidekoper SC, Fasotti L, Van Limbeek J. Recovery of standing balance in postacute stroke patients: a rehabilitation cohort study. Arch Phys Med Rehabil 2004;85:886–95. 10.1016/J.APMR.2003.05.012.

[120] Awad LN, Palmer JA, Pohlig RT, Binder-Macleod SA, Reisman DS. Walking speed and step length asymmetry modify the energy cost of walking after stroke. Neurorehabil Neural Repair 2015;29:416–23. 10.1177/1545968314552528.

[121] de Abreu DCC, Bandeira ACL, Magnani PE, de Oliveira Grigoletto DA, de Faria Junior JR, Teixeira VRS, et al. Standing balance test for fall prediction in older adults: a 6-month longitudinal study. BMC Geriatr 2024;24:1–9. 10.1186/S12877-024-05380-9/TABLES/4.

[122] Bauer CM, Gröger I, Rupprecht R, Marcar VL, Gaßmann KG. Prediction of future falls in a community dwelling older adult population using instrumented balance and gait analysis. Z Gerontol Geriatr 2016;49:232–6. 10.1007/S00391-015-0885-0.

[123] Horak FB, Nashner LM. Central programming of postural movements: adaptation to altered support-surface configurations. vol. 55. 1986.

[124] Loram ID, Lakie M. Direct measurement of human ankle stiffness during quiet standing: the intrinsic mechanical stiffness is insufficient for stability. J Physiol 2002;545:1041–53. 10.1113/JPHYSIOL.2002.025049.

[125] Gollnick PD, Sjödin B, Karlsson J, Jansson E, Saltin B. Human soleus muscle: A comparison of fiber composition and enzyme activities with other leg muscles. Pflugers Arch 1974;348:247–55. 10.1007/BF00587415/METRICS.

[126] Yamanaka E, Goto R, Kawakami M, Tateishi T, Kondo K, Nojima I. Intermuscular Coherence during Quiet Standing in Sub-Acute Patients after Stroke: An Exploratory Study. Brain Sciences 2023, Vol 13, Page 1640 2023;13:1640. 10.3390/BRAINSCI13121640.

[127] Bonan I V., Colle FM, Guichard JP, Vicaut E, Eisenfisz M, Tran Ba Huy P, et al. Reliance on visual information after stroke. Part I: Balance on dynamic posturography. Arch Phys Med Rehabil 2004;85:268–73. 10.1016/J.APMR.2003.06.017.

[128] Fimland MS, Moen PMR, Hill T, Gjellesvik TI, Tørhaug T, Helgerud J, et al. Neuromuscular performance of paretic versus non-paretic plantar flexors after stroke. Eur J Appl Physiol 2011;111:3041–9. 10.1007/S00421-011-1934-Z/TABLES/4.

[129] Peterson SM, Ferris DP. Differentiation in Theta and Beta Electrocortical Activity between Visual and Physical Perturbations to Walking and Standing Balance. ENeuro 2018;5. 10.1523/ENEURO.0207-18.2018.

130. Stokkermans M, Solis-Escalante T, Cohen MX, Weerdesteyn V. Distinct cortico-muscular coupling between step and stance leg during reactive stepping responses. 2023.

[131] Chen YC, Tsai YY, Chang GC, Hwang IS. Cortical reorganization to improve dynamic balance control with error amplification feedback. J Neuroeng Rehabil 2022;19:1–14. 10.1186/S12984-022-00980-1/FIGURES/6.

132. Marshall JL, Girgis FG, Zelko RR. The Biceps Femoris Tendon and Its Functional Significance. 1972.

[133] Patterson KK, Parafianowicz I, Danells CJ, Closson V, Verrier MC, Staines WR, et al. Gait Asymmetry in Community-Ambulating Stroke Survivors. Arch Phys Med Rehabil 2008;89:304–10. 10.1016/j.apmr.2007.08.142.

[134] Kumar S, Ferraro M, Nguyen L, Cao N, Ung N, Jose JS, et al. TMS assessment of corticospinal tract integrity after stroke: broadening the concept to inform neurorehabilitation prescription. Front Hum Neurosci 2024;18:1408818. 10.3389/FNHUM.2024.1408818/BIBTEX.

[135] Peng J, Zikereya T, Shao Z, Shi K. The neuromechanical of Beta-band corticomuscular coupling within the human motor system. Front Neurosci 2024;18:1441002. 10.3389/FNINS.2024.1441002/FULL.

[136] Davis NJ, Tomlinson SP, Morgan HM. The Role of Beta-Frequency Neural Oscillations in Motor Control. Journal of Neuroscience 2012;32:403–4. 10.1523/JNEUROSCI.5106-11.2012.

[137] Tzagarakis C, Ince NF, Leuthold AC, Pellizzer G. Beta-Band Activity during Motor Planning Reflects Response Uncertainty. Journal of Neuroscience 2010;30:11270–7. 10.1523/JNEUROSCI.6026-09.2010.

[138] Garnier YM, Lepers R, Canepa P, Martin A, Paizis C. Effect of the Knee and Hip Angles on Knee Extensor Torque: Neural, Architectural, and Mechanical Considerations. Front Physiol 2022;12:789867. 10.3389/FPHYS.2021.789867/BIBTEX.

[139] Mima T, Steger JÈ, Schulman AE, Gerloff C, Hallett M. Electroencephalographic measurement of motor cortex control of muscle activity in humans. 2000.

[140] Gao Z, Lv S, Ran X, Wang Y, Xia M, Wang J, et al. Influencing factors of corticomuscular coherence in stroke patients. Front Hum Neurosci 2024;18:1354332. 10.3389/FNHUM.2024.1354332/XML.

[141] Schulz R, Bönstrup M, Guder S, Liu J, Frey B, Quandt F, et al. Corticospinal Tract Microstructure Correlates With Beta Oscillatory Activity in the Primary Motor Cortex After Stroke. Stroke 2021;52:3839–47. 10.1161/STROKEAHA.121.034344.

[142] Barone J, Rossiter HE. Understanding the Role of Sensorimotor Beta Oscillations. Front Syst Neurosci 2021;15:655886. 10.3389/FNSYS.2021.655886/BIBTEX.

[143] Schmidt R, Ruiz MH, Kilavik BE, Lundqvist M, Starr PA, Aron AR. Beta Oscillations in Working Memory, Executive Control of Movement and Thought, and Sensorimotor Function. Journal of Neuroscience 2019;39:8231–8. 10.1523/JNEUROSCI.1163-19.2019.

[144] Fujita K, Tsushima Y, Hayashi K, Kawabata K, Ogawa T, Hori H, et al. Altered muscle synergy structure in patients with poststroke stiff knee gait. Sci Rep 2024;14. 10.1038/S41598-024-71083-1.

[145] Sui J, Adali T, Yu Q, Chen J, Calhoun VD. A review of multivariate methods for multimodal fusion of brain imaging data. J Neurosci Methods 2012;204:68–81. 10.1016/j.jneumeth.2011.10.031.

[146] Arsenault D, Ivanova TD, Garland SJ. Postural control in response to unilateral and bilateral external perturbations in older adults. Gait Posture 2022;94:26–31. 10.1016/J.GAITPOST.2022.02.024.

[147] Lan N, Funato T, Jadlovska S, Morasso P. Integrating ankle and hip strategies for the stabilization of upright standing: An intermittent control model. Front Comput Neurosci 2022;16:956932. 10.3389/FNCOM.2022.956932.

[148] Blenkinsop GM, Pain MTG, Hiley MJ. Balance control strategies during perturbed and unperturbed balance in standing and handstand. R Soc Open Sci 2017;4:161018. 10.1098/RSOS.161018.

